# The statistical analysis of daily data associated with different parameters of the New Coronavirus COVID-19 pandemic in Georgia and their two-week interval prediction in summer 2021

**DOI:** 10.1101/2021.09.08.21263265

**Authors:** Avtandil G. Amiranashvili, Ketevan R. Khazaradze, Nino D. Japaridze

## Abstract

The lockdown introduced in Georgia on November 28, 2020 brought positive results. There were clearly positive tendencies in the spread of COVID-19 to February - first half of March 2021. However, in April-May 2021 there was a significant deterioration in the epidemiological situation. From June to August 2021, the epidemiological situation with Covid-19 in Georgia became very difficult.

In this work results of the next statistical analysis of the daily data associated with New Coronavirus COVID-19 infection of confirmed (C), recovered (R), deaths (D) and infection rate (I) cases of the population of Georgia in the period from June 01, 2021 to August 31, 2021 are presented. It also presents the results of the analysis of two-week forecasting of the values of C, D and I. As earlier, the information was regularly sent to the National Center for Disease Control & Public Health of Georgia and posted on the Facebook page https://www.facebook.com/Avtandil1948/.

The analysis of data is carried out with the use of the standard statistical analysis methods of random events and methods of mathematical statistics for the non-accidental time-series of observations. In particular, the following results were obtained.

Georgia’s ranking in the world for Covid-19 monthly mean values of infection and deaths cases in summer 2021 (per 1 million population) was determined. Among 159 countries with population ≥ 1 million inhabitants in August 2021 Georgia was in the 1 place on new infection cases and on Death.

A comparison between the daily mortality from Covid-19 in Georgia in summer 2021 with the average daily mortality rate in 2015-2019 shows, that the largest share value of D from mean death in 2015-2019 was 66.0 % (26.08.2021 and 31.08.2021), the smallest 6.0 % (09.07.2021).

The statistical analysis of the daily and decade data associated with coronavirus COVID-19 pandemic of confirmed, recovered, deaths cases and infection rate of the population of Georgia are carried out. Maximum daily values of investigation parameters are following: C = 6208 (17.08.2021), R = 6177 (29.08.2021), D = 79 (26.08.2021 and 31.08.2021), I = 13.0 % (17.08.2021). Maximum mean decade values of investigation parameters are following: C = 5019 (2 Decade of August 2021), R = 4822 (3 Decade of August 2021), D = 69 (3 Decade of August 2021), I = 10.88 % (2 Decade of August 2021).

It was found that as with September 2020 to February 2021 and in spring 2021 [7,8], from June to August 2021 the regression equations for the time variability of the daily values of C, R and D have the form of a tenth order polynomial.

Mean values of speed of change of confirmed -V(C), recovered - V(R), deaths - V(D) and infection rate V(I) coronavirus-related cases in different decades of months in the summer 2021 were determined. Maximum mean decade values of investigation parameters are following: V(C) = +134 cases/day (1 Decade of August 2021), V(R) = +134 cases/day (2 Decade of August 2021), V(D) = +2.4 cases/day (3 Decade of August 2021), V(I) = + 0.25 %/ day (1 decades of August 2021).

Cross-correlations analysis between confirmed COVID-19 cases with recovered and deaths cases shows, that the maximum effect of recovery is observed 19 days after infection (RC=0.95), and deaths - after 16 and 18 days (RC=0.94). In Georgia in the summer 2021, the duration of the impact of the delta variant of the coronavirus on people (recovery, mortality) could be up to two months.

Comparison of real and calculated predictions data of C, D and I in Georgia are carried out. It was found that in summer 2021 two-week daily and mean two-week real values of C, D and I practically fall into the 67% - 99.99% confidence interval of these predicted values.

With September 1, 2021, it is started monthly forecasting of C, D and I values.

As earlier, the comparison of data about C and D in Georgia (GEO) with similar data in Armenia (ARM), Azerbaijan (AZE), Russia (RUS), Turkey (TUR) and in the World (WRL) is also carried out.

## 1. Introduction

Twenty months has passed since the outbreak of the novel coronavirus (COVID-19) in China, recognized on March 11, 2020 as a pandemic due to its rapid spread in the world [1]. Over the indicated period of time, despite the measures taken (including vaccination), the overall level of morbidity and mortality in many countries of the world remains quite high. Together with epidemiologists, scientists and specialists of various disciplines from all over the world continue intensive research of this unprecedented phenomenon (including Georgia [2-8]).

So in our works [5-8] it was noted that specialists in the field of physical and mathematical sciences make an important contribution to research on the spread of the new coronavirus COVID-19. Work on systematization, statistical analysis, forecasting, spatial-temporal modeling of the spread of the new coronavirus is being actively continued at the present time [9-19, etc.].

This work is a continuation of the researches [5-8].

In this work results of a statistical analysis of the daily data associated with New Coronavirus COVID-19 infection of confirmed (C), recovered (R), deaths (D) and infection rate (I) cases of the population of Georgia in the period from June 01, 2021 to August, 2021 are presented. It also presents the results of the analysis of two-week forecasting of the values of C, D and I. The information was regularly sent to the National Center for Disease Control & Public Health of Georgia and posted on the Facebook page https://www.facebook.com/Avtandil1948/.

The comparison of data about C and D in Georgia with similar data in Armenia, Azerbaijan, Russia, Turkey and in the world is also carried out.

We used standard methods of statistical analysis of random events and methods of mathematical statistics for non-random time series of observations [6-8, 20, 21].

## 2. Study areas, material and methods

The study area: Georgia. Data of John Hopkins COVID-19 Time Series Historical Data (with US State and County data) [https://www.soothsawyer.com/john-hopkins-time-series-data-with-us-state-and-county-city-detail-historical/; https://data.humdata.org/dataset/total-covid-19-tests-performed-by-country] and https://stopcov.ge about daily values of confirmed, recovered, deaths and infection rate coronavirus-related cases, from June 01, 2021 to August 31, 2021 are used. The work also used data of National Statistics Office of Georgia (Geostat) on the average monthly total mortality in Georgia in October-May 2015-2019 [https://www.geostat.ge/en/]. In the proposed work the analysis of data is carried out with the use of the standard statistical analysis methods of random events and methods of mathematical statistics for the non-accidental time-series of observations [6-8, 20, 21].

The following designations will be used below: Mean – average values; Min – minimal values; Max - maximal values; Range – Max-Min; St Dev - standard deviation; σ_m_ - standard error; CV = 100□St Dev/Mean – coefficient of variation, %; R^2^ – coefficient of determination; r – coefficient of linear correlation; CR – coefficient of cross correlation; Lag = 1, 2…30 Day; K_DW_ – Durbin-Watson statistic; Calc – calculated data; Real - measured data; D1, D2, D3 – numbers of the month decades; α - the level of significance; C, R, D - daily values of confirmed, recovered and deaths coronavirus-related cases; V(C), V(R) and V(D) - daily values of speed of change of confirmed, recovered and deaths coronavirus-related cases (cases/day); I - daily values of infection rate (or positive rate) coronavirus-related cases (100□ C/number of coronavirus tests performed); V(I) - daily values of speed of change of I; DC – deaths coefficient, % = (100□D/C). Official data on number of coronavirus tests performed are published from December 05, 2020 [https://stopcov.ge].

The statistical programs Data Fit 7, Mesosaur and Excel 16 were used for calculations.

The curve of trend is equation of the regression of the connection of the investigated parameter with the time at the significant value of the determination coefficient and such values of K_DW_, where the residual values are accidental. If the residual values are not accidental the connection of the investigated parameter with the time we will consider as simply regression.

The calculation of the interval prognostic values of C, D and I taking into account the periodicity in the time-series of observations was carried out using Excel 16 (the calculate methodology was description in [6]). The duration of time series of observations for calculating two-week forecasts of the values of C, D and I was 35 - 74 days.

67%…99.99%_Low - 67% 99.99% lower level of confidence interval of prediction values of C, D and I; 67%…99.99%_Upp - 67% 99.99% upper level of confidence interval of prediction values of C, D and I.

In the Table 1 [6] the scale of comparing real data with the predicted ones and assessing the stability of the time series of observations in the forecast period in relation to the pre-predicted one (period for prediction calculating) is presented.

**Table 1.** The statistical characteristics of Covid-19 confirmed cases to 1 million populations in Georgia, neighboring countries (Armenia, Azerbaijan, Russia, Turkey) and World from June 1, 2021 to August 31, 2021 (r_min_ = ± 0.21, α = 0.05).

| Parameter | GEO | ARM | AZE | RUS | TUR | WRL |
| --- | --- | --- | --- | --- | --- | --- |
| Mean | 600 | 71 | 97 | 135 | 146 | 65 |
| Min | 70 | 9 | 2 | 60 | 0 | 38 |
| Max | 1665 | 233 | 415 | 173 | 546 | 110 |
| Range | 1595 | 224 | 413 | 113 | 546 | 72 |
| Median | 410 | 50 | 18 | 144 | 84 | 60 |
| St Dev | 460 | 54 | 134 | 32 | 100 | 18 |
| $\sigma_m$ | 48.2 | 5.7 | 14.1 | 3.4 | 10.4 | 1.9 |
| Cv (%) | 76.6 | 75.9 | 138.8 | 23.7 | 68.4 | 27.7 |
| **Correlation Matrix** |  |  |  |  |  |  |
| GEO | 1 | 0.88 | 0.81 | 0.27 | 0.75 | 0.76 |
| ARM | 0.88 | 1 | 0.92 | 0.21 | 0.63 | 0.72 |
| AZE | 0.81 | 0.92 | 1 | 0.07 | 0.61 | 0.62 |
| RUS | 0.27 | 0.21 | 0.07 | 1 | 0.22 | 0.31 |
| TUR | 0.75 | 0.63 | 0.61 | 0.22 | 1 | 0.74 |
| WRL | 0.76 | 0.72 | 0.62 | 0.31 | 0.74 | 1 |

## 3. Results and Discussion

The results in the Fig. 1-24 and Table 1-9 are presented.

**Fig. 1.**
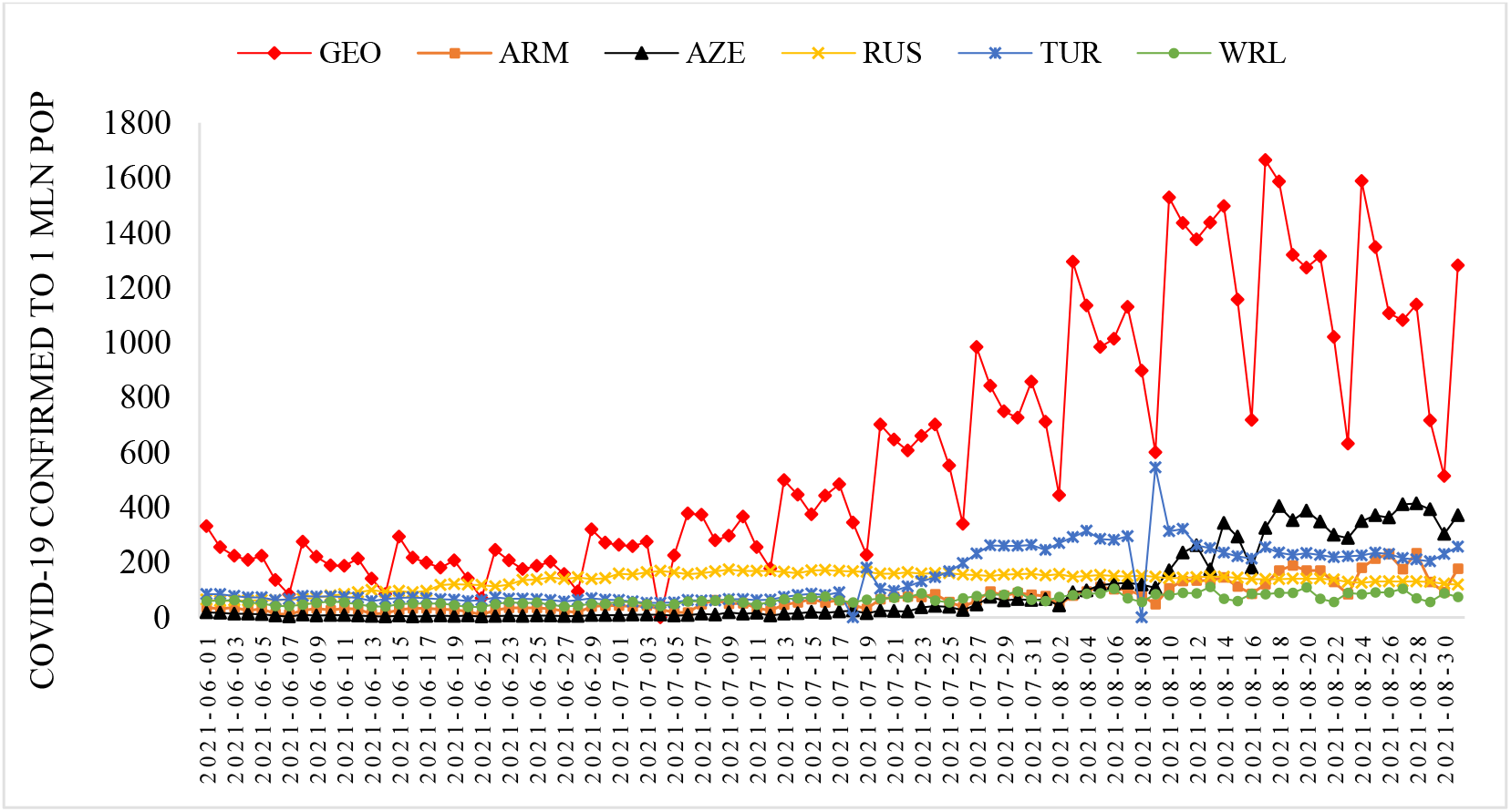
Time-series of Covid-19 confirmed cases to 1 million populations in Georgia, neighboring countries (Armenia, Azerbaijan, Russia, Turkey) and World from June 1, 2021 to August 31, 2021.

### 3.1 Comparison of time-series of Covid-19 confirmed and deaths cases in Georgia, its neighboring countries and World in summer 2021

The time-series curves and statistical characteristics of Covid-19 confirmed and deaths cases (to 1 million populations) in Georgia, its neighboring countries and World from June 01, 2021 to August 31, 2021 in Fig. 1 - 2 and in Table 1-2 are presented.

Variability of the values of C per 1 million populations is as follows (Fig. 1, Table 1):

- Georgia. Range of change: 70-1665, mean value - 600.
- Armenia. Range of change: 9-233, mean value - 71.
- Azerbaijan. Range of change: 2-415, mean value - 97.
- Russia. Range of change: 60-173, mean value - 135.
- Turkey. Range of change: 0-546, mean value -146.
- World. Range of change: 38-110, mean value - 65.

The largest variations in C values were observed in Azerbaijan (C_V_=138.8%), the smallest - in Russia (C_V_=23.7 %).

Significant linear correlation (rmin = ± 0.21, α = 0.05) between these countries on C value varies from 0.21 (pair Armenia-Russia) to 0.92 (pair Armenia-Azerbaijan). Linear correlation between World and these countries is significant and varied from 0.31 (pair World-Russia) to 0.76 (pair World-Georgia). Thus, in summer 2021 in these countries there were more or less similar trends to the world the development of the coronavirus epidemic (Table 1).

Variability of the values of D per 1 million populations is as follows (Fig. 2, Table 2):

**Table 2.**
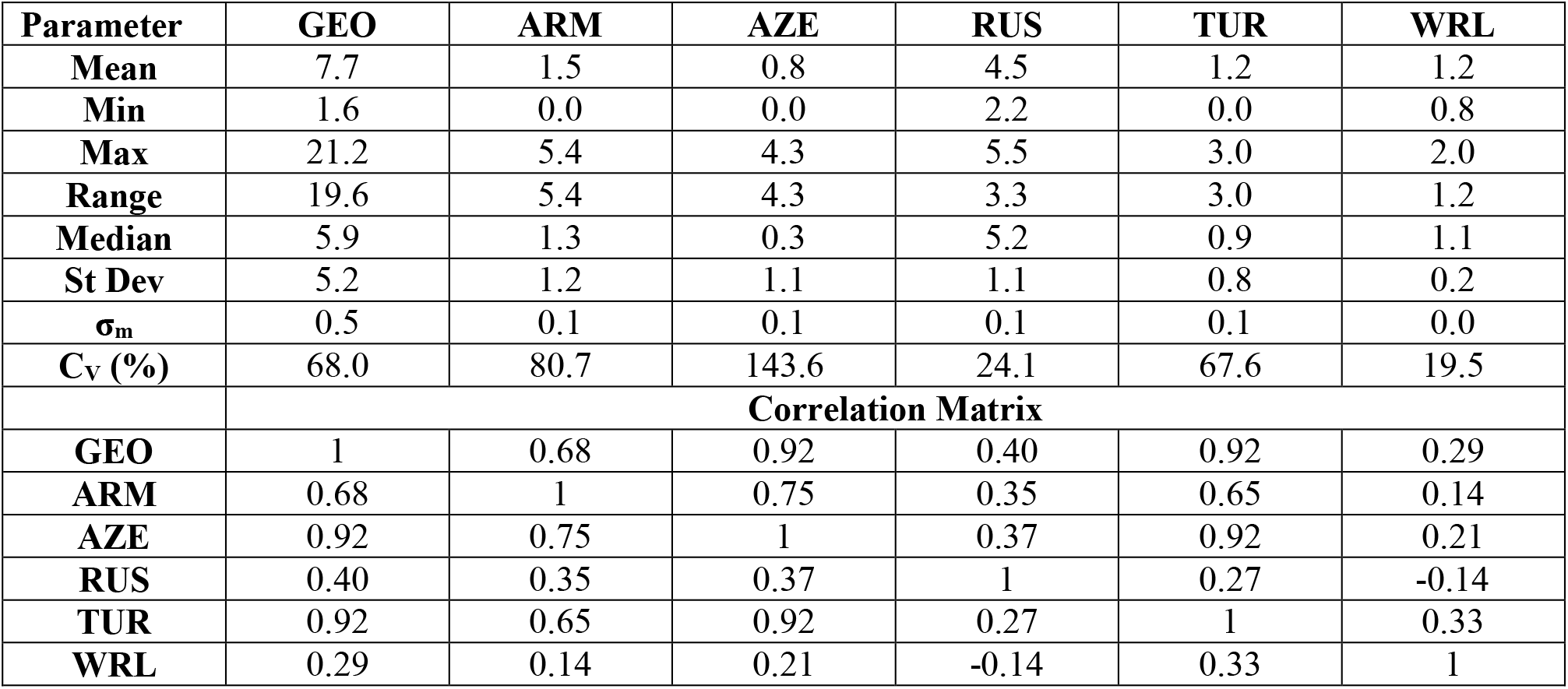
The statistical of deaths cases from Covid-19 to 1 million populations in Georgia, neighboring countries (Armenia, Azerbaijan, Russia, Turkey) and World from June 1, 2021 to August 31, 2021. (r_min_ = ± 0.21, α = 0.05).

**Fig. 2.**
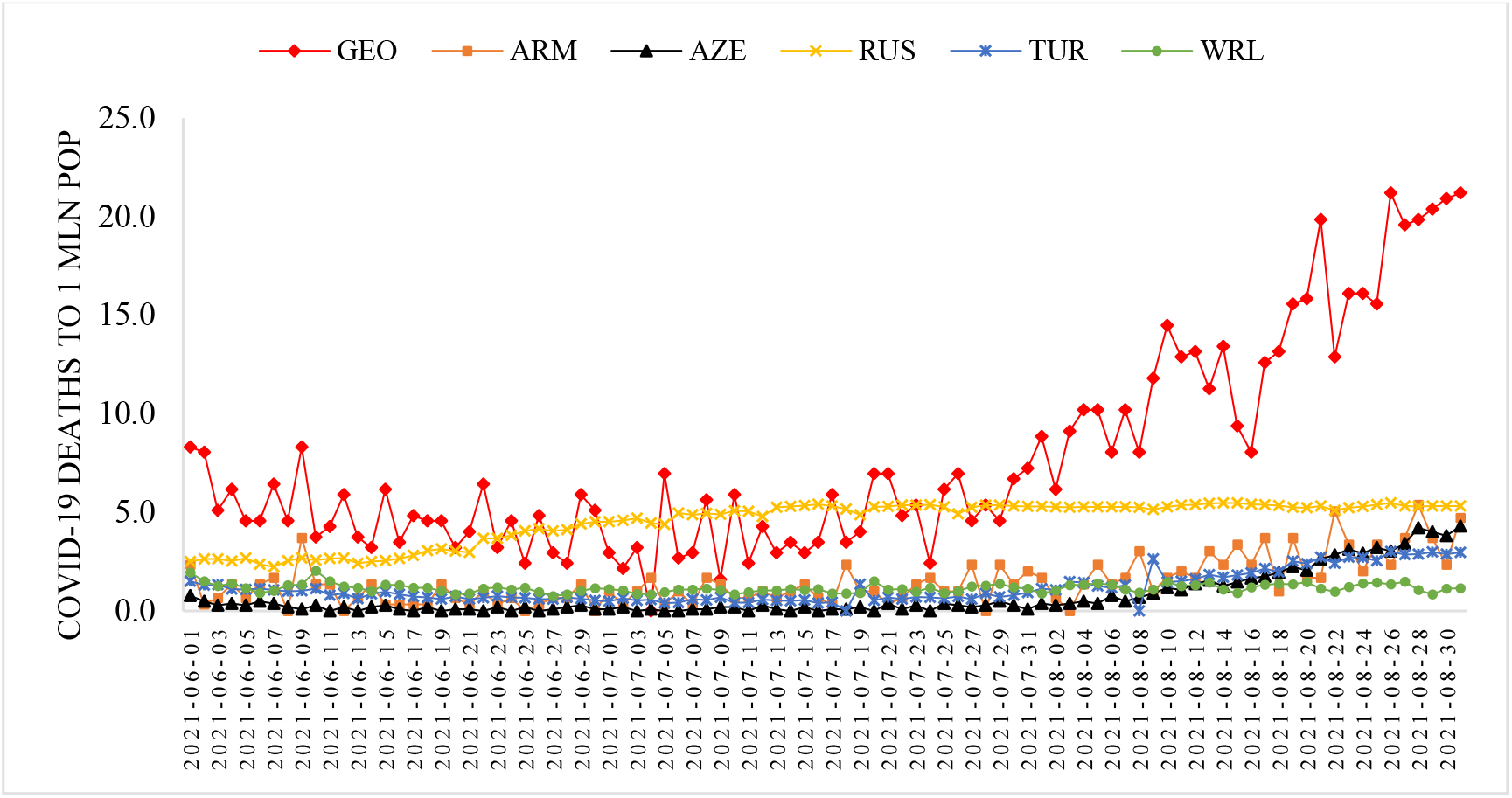
Time-series of deaths cases from Covid-19 to 1 million population in in Georgia, neighboring countries (Armenia, Azerbaijan, Russia, Turkey) and World from June 1, 2021 to August 31, 2021.

- Georgia. Range of change: 1.6-21.2, mean value – 7.7.
- Armenia. Range of change: 0.0-5.4, mean value – 1.5.
- Azerbaijan. Range of change: 0.0-4.3, mean value – 0.8.
- Russia. Range of change: 2.2-5.5, mean value – 4.5.
- Turkey. Range of change: 0.0-3.0, mean value -1.2.
- World. Range of change: 0.8-2.0, mean value – 1.2.

The largest variations in D values were observed in Azerbaijan (C_V_=143.6%), the smallest - in Russia (C_V_=24.1 %).

Significant linear correlation between these countries on D value varies from 0.27 (pair Russia-Turkey) to 0.92 (pair Georgia-Azerbaijan and pair Georgia - Turkey). Linear correlation between World and these countries besides Armenia and Russia is significant and varied from 0.21 (pair World-Azerbaijan) to 0.33 (pair World-Turkey). Thus, for Georgia, Azerbaijan and Turkey in summer 2021 there were some similar trends with world in the variability of mortality from coronavirus (Table 2).

In Table 3 the statistical characteristics of mean monthly values of C and D related to Covid-19 for 159 countries with population ≥ 1 million inhabitants and World in the summer 2021 (normed per 1 million population) is presented.

**Table 3.** The statistical characteristics of mean monthly values of confirmed cases and Deaths cases related to Covid-19 for 159 countries with population ≥ 1 million inhabitant and World from June to August 2021 (normed on 1 mln pop.).

| Month | June |  | July |  | August |  |
| --- | --- | --- | --- | --- | --- | --- |
| Parameter | C | D | C | D | C | D |
| Mean | 70 | 1.4 | 87 | 1.3 | 120 | 1.5 |
| Min | 0 | 0 | 0 | 0 | 0 | 0.0 |
| Max | 716 | 17.1 | 552 | 19.0 | 1127 | 13.7 |
| Range | 716 | 17.1 | 552 | 19.0 | 1127 | 13.7 |
| St Dev | 121 | 2.7 | 123 | 2.4 | 170 | 2.3 |
| $\sigma_m$ | 10 | 0.2 | 10 | 0.2 | 14 | 0.2 |
| Cv (%) | 174 | 189 | 141 | 189 | 142 | 149 |

As follows from this Table range of change of mean monthly values of C for 159 countries varied from 0 (all months) to 1127 (August). Average value of C for 159 countries varied from 70 (June) to 120 (August). Value of C_V_ changes from 141% (July) to 174% (June).

Range of change of mean monthly values of D for 159 countries varied from 0 (all months) to 19.0 (July). Average value of D for 159 countries varied from 1.3 (July) to 1.5 (August). Value of C_V_ changes from 149% (August) to 189% (June and July).

Between mean monthly values of Deaths and Confirmed cases related to Covid-19 for 159 countries with population ≥ 1 million inhabitants and World linear correlation and regression are observed (Fig. 3). As follows from Fig. 3 the highest growth rate of the monthly average values of D depending on C was observed in June, the smallest - in August (the corresponding values of the linear regression coefficients).

**Fig. 3.**
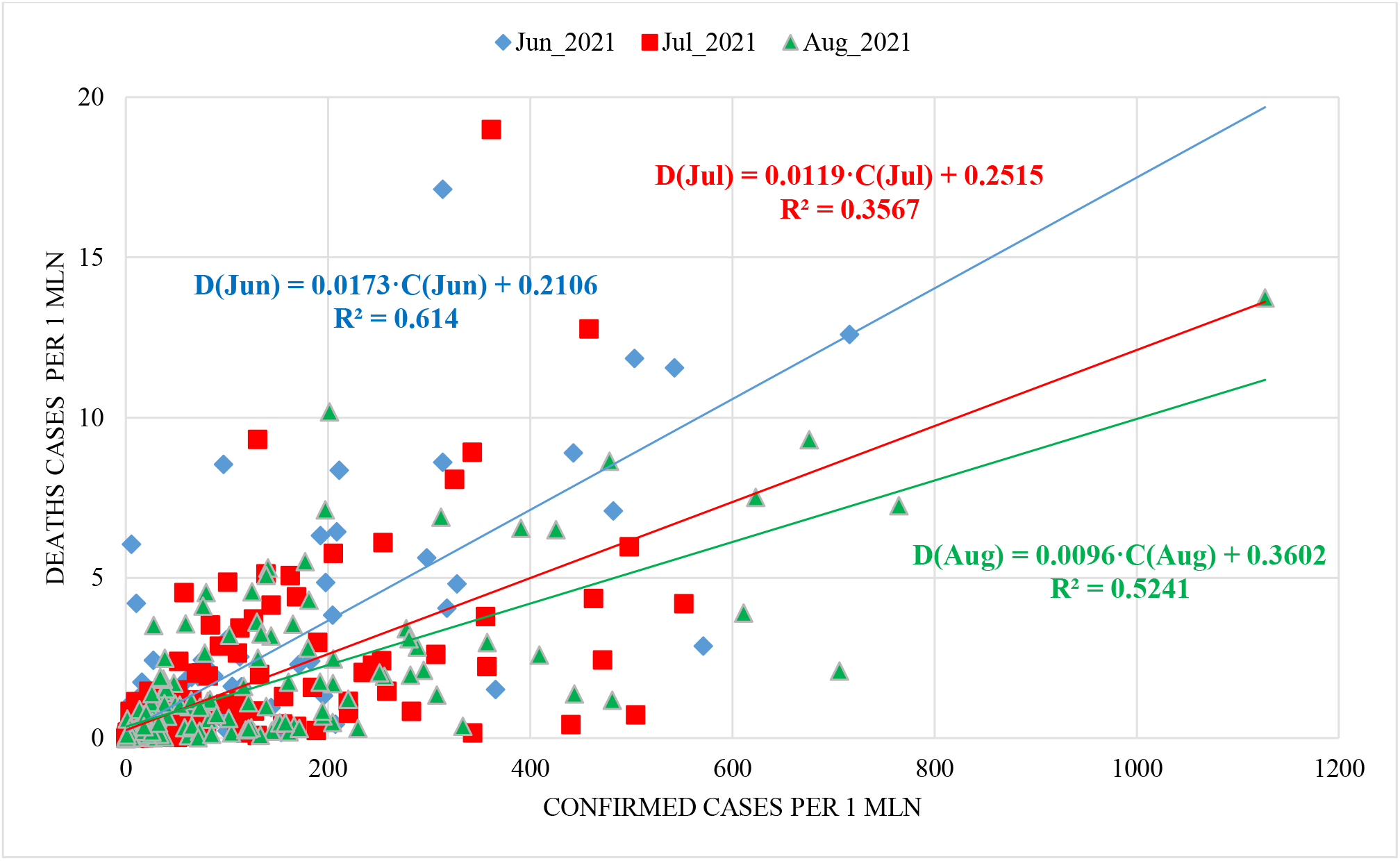
Linear correlation and regression between mean monthly values of Deaths and Confirmed cases related to Covid-19 for 159 countries with population ≥ 1 million inhabitants and world (normed on 1 mln pop.).

In Table 4 data about Covid-19 mean monthly values of infection (C) and deaths (D) cases in summer 2021 (per 1 million population) and ranking of Georgia, Armenia, Azerbaijan, Russia and Turkey by these parameters (in brackets) among 159 countries with population ≥ 1 million inhabitant are presented. The corresponding values of the deaths coefficient (DC) are also given here.

**Table 4.**
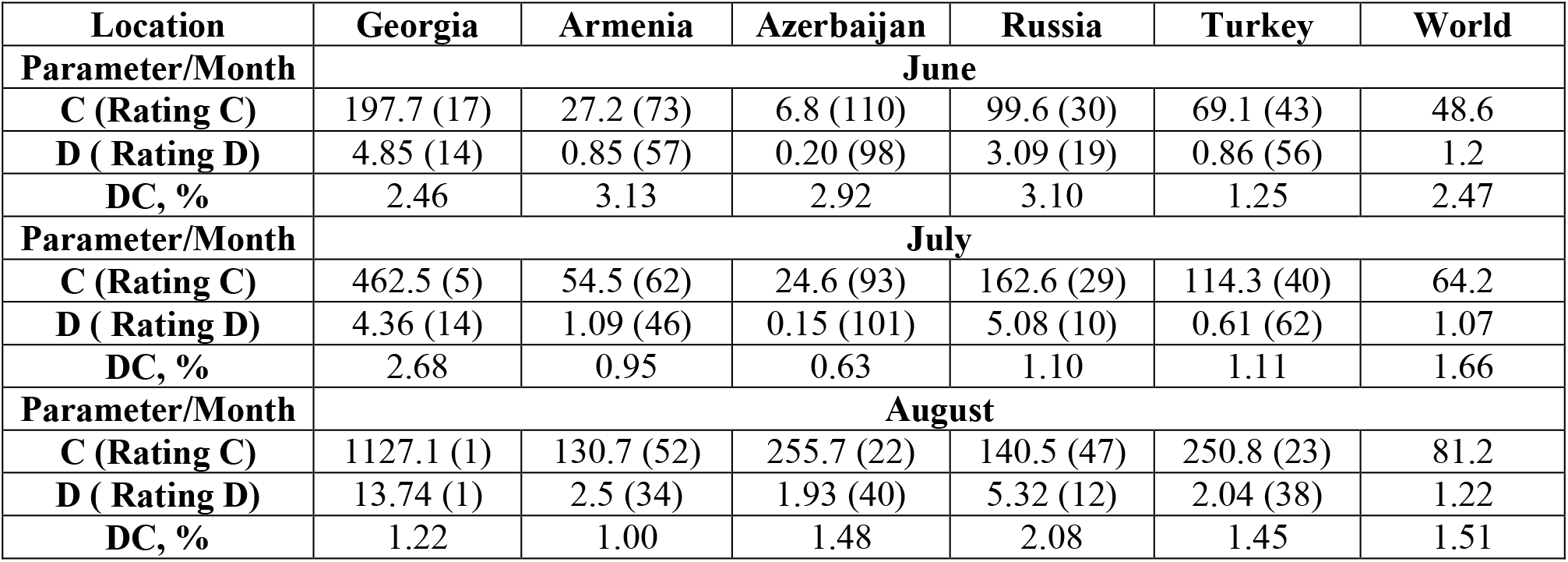
Covid-19 mean monthly values of infection (C) and deaths (D) cases in summer 2021 (per 1 million population) and ranking of Georgia, Armenia, Azerbaijan, Russia, Turkey by these parameters (in brackets) among 159 countries with population ≥ 1 million inhabitant, and correspondent values of deaths coefficient (DC).

| Location | Georgia | Armenia | Azerbaijan | Russia | Turkey | World |
| --- | --- | --- | --- | --- | --- | --- |
| **Parameter/Month** | **June** |  |  |  |  |  |
| **C (Rating C)** | 197.7 (17) | 27.2 (73) | 6.8 (110) | 99.6 (30) | 69.1 (43) | 48.6 |
| **D ( Rating D)** | 4.85 (14) | 0.85 (57) | 0.20 (98) | 3.09 (19) | 0.86 (56) | 1.2 |
| **DC, %** | 2.46 | 3.13 | 2.92 | 3.10 | 1.25 | 2.47 |
| **Parameter/Month** | **July** |  |  |  |  |  |
| **C (Rating C)** | 462.5 (5) | 54.5 (62) | 24.6 (93) | 162.6 (29) | 114.3 (40) | 64.2 |
| **D ( Rating D)** | 4.36 (14) | 1.09 (46) | 0.15 (101) | 5.08 (10) | 0.61 (62) | 1.07 |
| **DC, %** | 2.68 | 0.95 | 0.63 | 1.10 | 1.11 | 1.66 |
| **Parameter/Month** | **August** |  |  |  |  |  |
| **C (Rating C)** | 1127.1 (1) | 130.7 (52) | 255.7 (22) | 140.5 (47) | 250.8 (23) | 81.2 |
| **D ( Rating D)** | 13.74 (1) | 2.5 (34) | 1.93 (40) | 5.32 (12) | 2.04 (38) | 1.22 |
| **DC, %** | 1.22 | 1.00 | 1.48 | 2.08 | 1.45 | 1.51 |

In particular, as follows from this Table, mean monthly values of C for 5 country changes from 6.8 (Azerbaijan, June, 110 place between 159 country) to 1127.1 (Georgia, August, 1 place between 159 country).

Mean monthly values of D for 5 country changes from 0.15 (Azerbaijan, July, 101 place between 159 country) to 13.74 (Georgia, August, 1 place between 159 country).

So, among 159 countries with population ≥ 1 million inhabitants and World in August 2021 Georgia was in the 1 place on new infection cases and Death.

Mean monthly values of DC for 5 country changes from 0.63% (Azerbaijan, July) to 3.10 (Russia, June).

Note that the mean values of DC (%) in summer 2021 are: Georgia – 1.28, Armenia – 2.08, Azerbaijan

– 0.80, Russia – 3.36, Turkey – 0.80, World – 1.79 (according to data from Table 1 and 2).

### 3.2 Comparison of daily death from Covid-19 in Georgia with daily mean death in 2015-2019 from June 1, 2021 to August 31, 2021

In Fig. 4-7 data of the daily death from Covid-19 in Georgia from October 1, 2020 to August 31, 2021 in comparison with daily mean death in 2015-2019. The daily mean death in 2015-2019 in different months are: October - 123, November - 135, December - 144, January - 155, February – 146, March - 141, April - 137, May – 131, June – 124, July – 117, August – 120 (Fig. 4, 6).

**Fig. 4.**
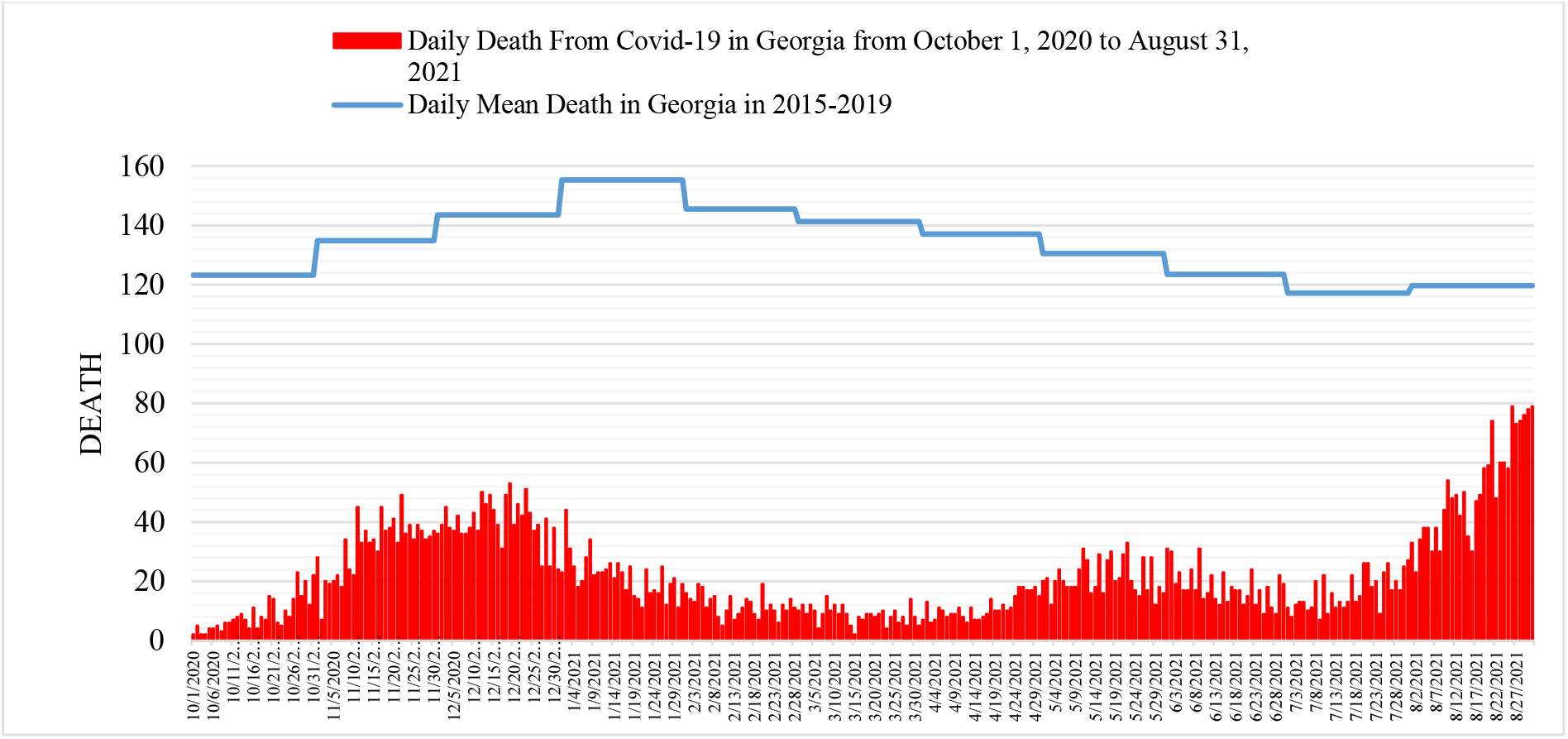
Daily death from Covid-19 in Georgia from October 1, 2020 to August 31, 2021 in comparison with daily mean death in 2015-2019.

**Fig. 5.**
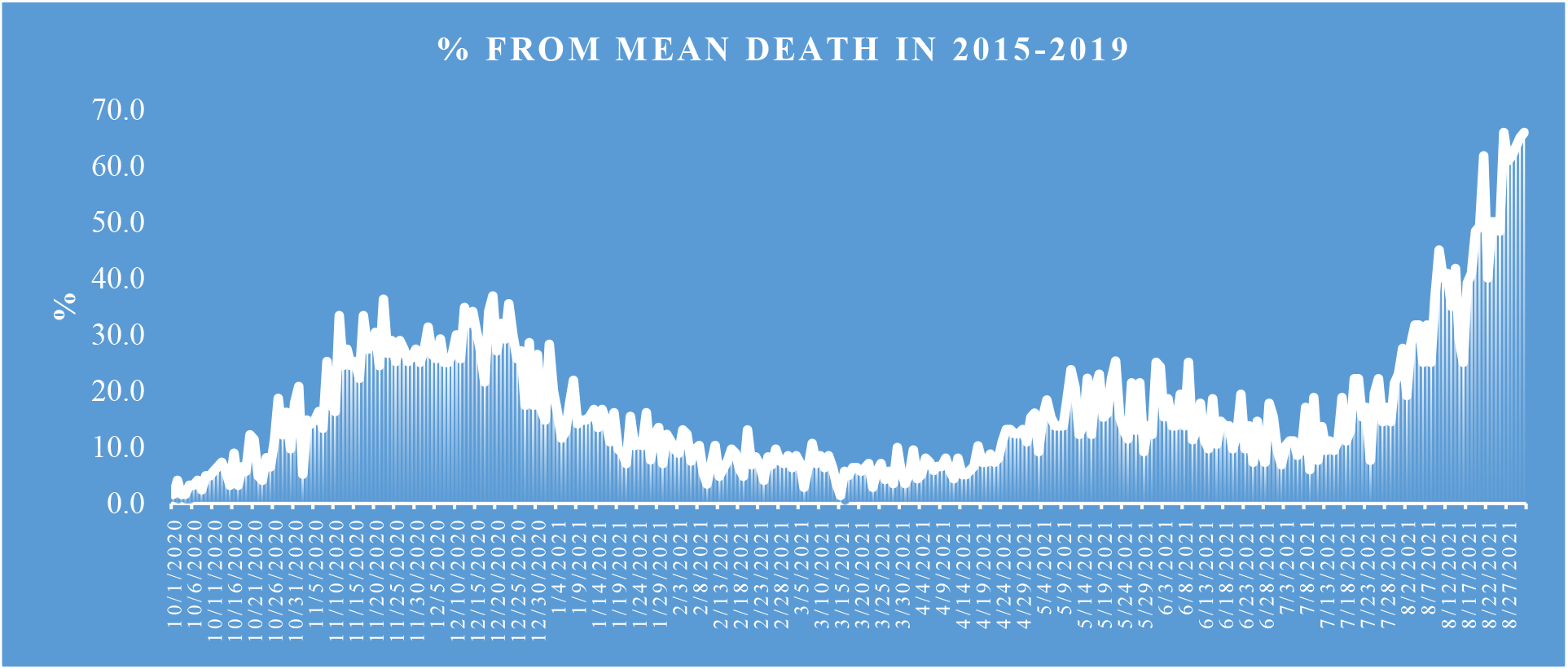
Share value of D from mean death in 2015-2019.

**Fig. 6.**
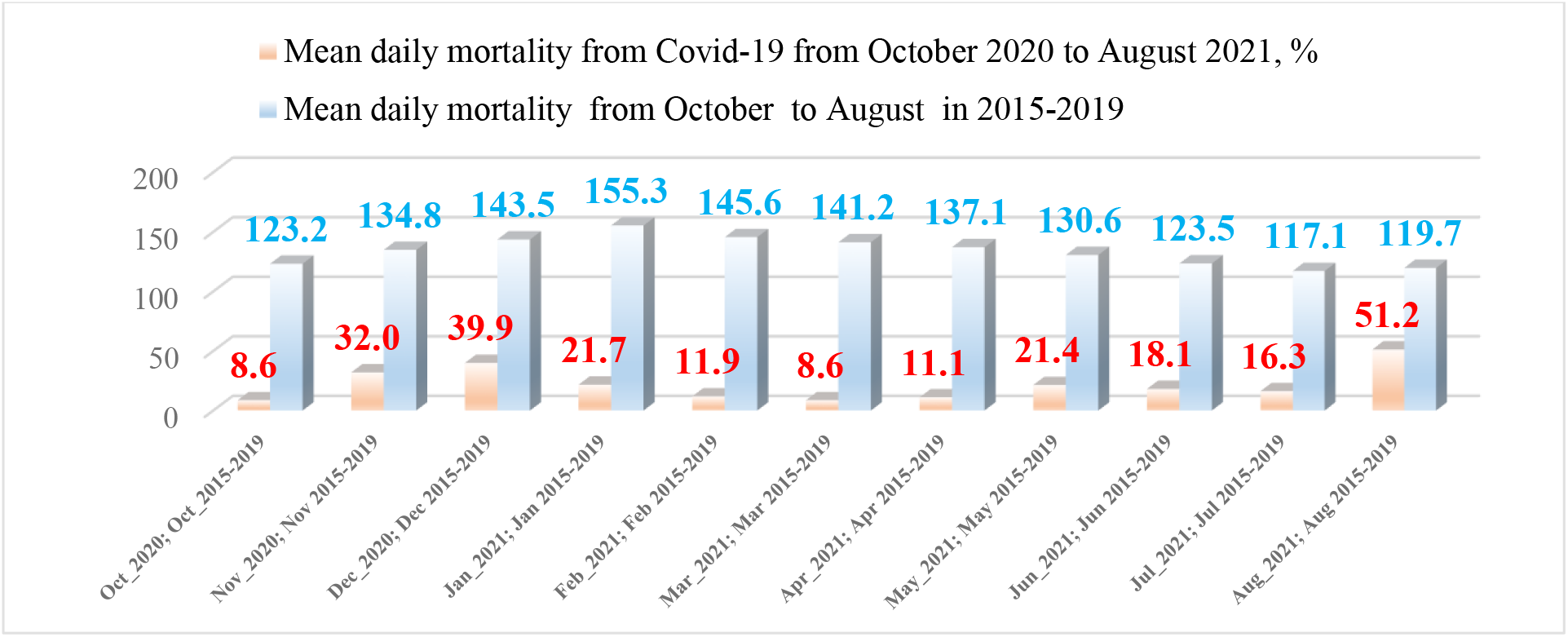
Mean daily mortality from Covid-19 from October 2020 to August 2021, % and mean daily mortality from October to August in 2015-2019

The daily death from Covid-19 from June 1, 2021 to August 31, 2021 changes from 7 to 79 (Fig. 4). A comparison between the daily mortality from Covid-19 in Georgia in summer 2021 with the average daily mortality rate in 2015-2019 shows (Fig. 5), that the largest share value of D from mean death in 2015-2019 was 66.0 % (26.08.2021 and 31.08.2021), the smallest 6.0 % (09.07.2021).

The most share of mean daily mortality from Covid-19 of mean daily mortality in 2015-2019 from October 2020 to August 2021 in August was observed – 42.8% (Fig. 7).

**Fig. 7.**
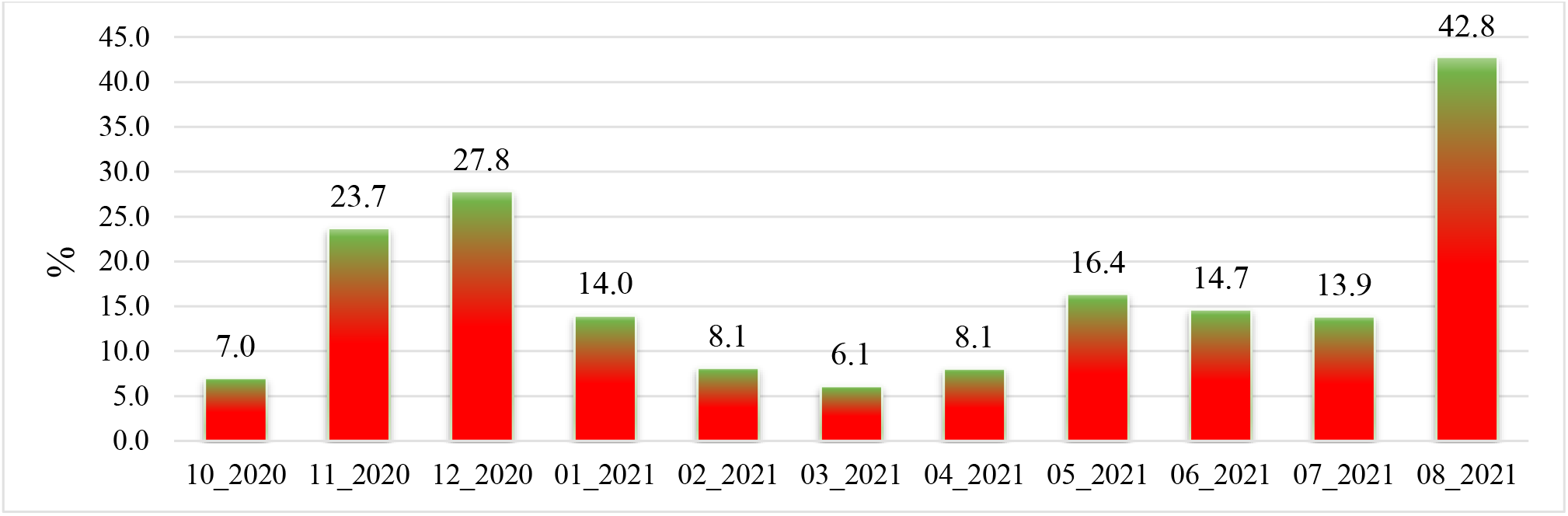
Share of mean daily mortality from Covid-19 of mean daily mortality in 2015-2019 from October 2020 to August 2021.

### 3.3 The statistical analysis of the daily and decade data associated with New Coronavirus COVID-19 pandemic in summer 2021

Results of the statistical analysis of the daily and decade data associated with New Coronavirus COVID-19 pandemic in Georgia from June 1, 2021 to August 31, 2021 in Tables 5-7 and Fig. 8 – 20 are presented.

**Table 5.** Statistical characteristics of the daily data associated with coronavirus COVID-19 pandemic of confirmed, recovered, deaths cases and infection rate of the population of Georgia from 01.06.2021 to 31.08.2021.

| Parameter | Confirmed | Recovered | Deaths | Infection Rate, % |
| --- | --- | --- | --- | --- |
| Mean | 2237 | 1791 | 29 | 6.1 |
| Min | 260 | 361 | 7 | 1.8 |
| Max | 6208 | 6177 | 79 | 13.0 |
| Range | 5948 | 5816 | 72 | 11.1 |
| St Dev | 1714 | 1513 | 19 | 3.4 |
| $\sigma_m$ | 180 | 159 | 2.0 | 0.4 |
| $C_v$ (%) | 76.6 | 84.5 | 68.0 | 55.3 |

**Fig. 8.**
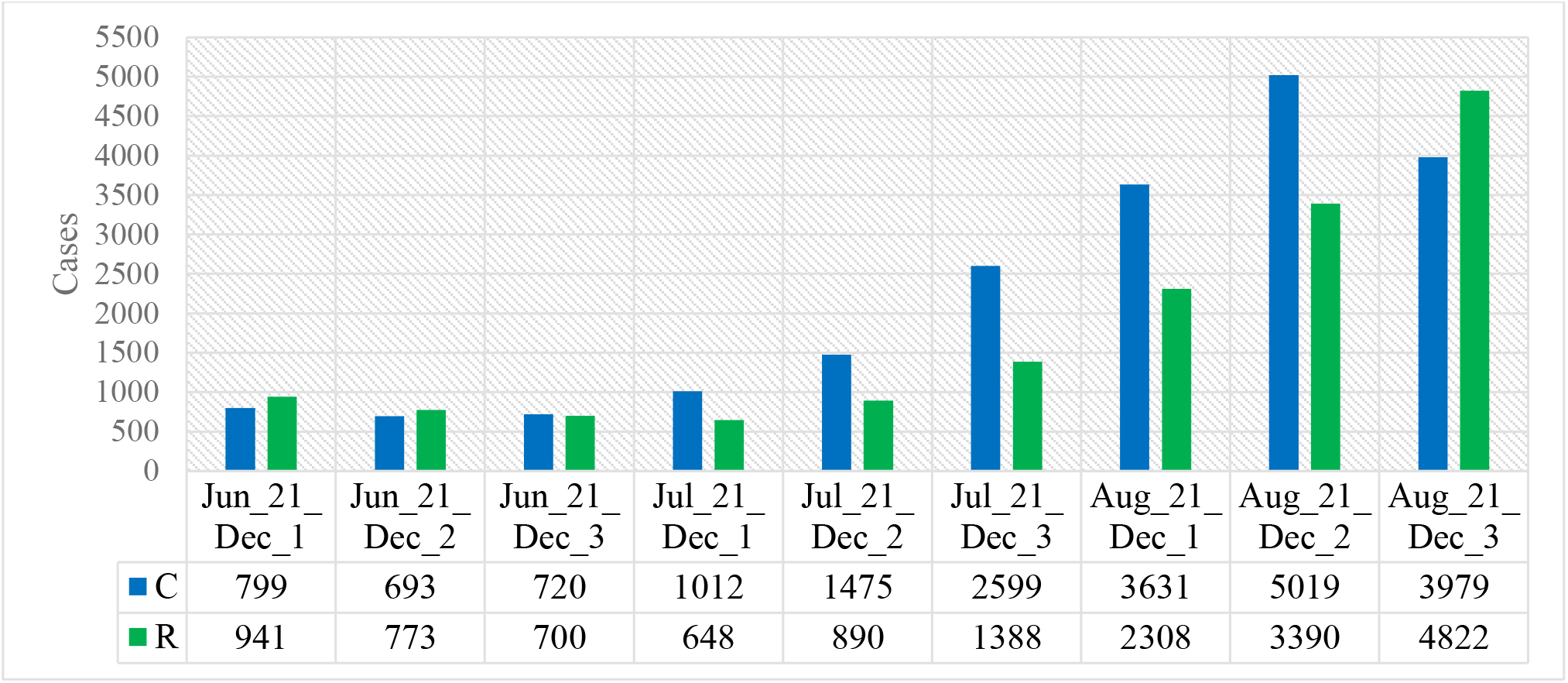
Mean values of confirmed and recovered coronavirus-related cases in different decades of months in Georgia from June 2021 to August 2021.

The mean and extreme values of the studied parameters are as follows (Table 5): C - mean - 2237, range: 260 - 6208; R - mean - 1791, range: 361 - 6177; D - mean - 29, range: 7 - 79; I (%) - mean – 6.1, range: 1.8 – 13.0. All studied parameters are subject to notable variations: 55.3 % (I) ≤C_**V**_ ≤84.5% (R).

Mean decade values of confirmed and recovered coronavirus-related cases varies within the following limits (Fig. 8): C – from 693 (2 Decade of June 2021) to 5019 (2 Decade of August 2021); R – from 648 (1 Decade of July 2021) to 4822 (3 Decade of August 2021).

Mean decade values of deaths coronavirus-related cases (Fig. 9) varies from 13 (1 Decades of July) to 69 (3 Decade of August).

**Fig. 9.**
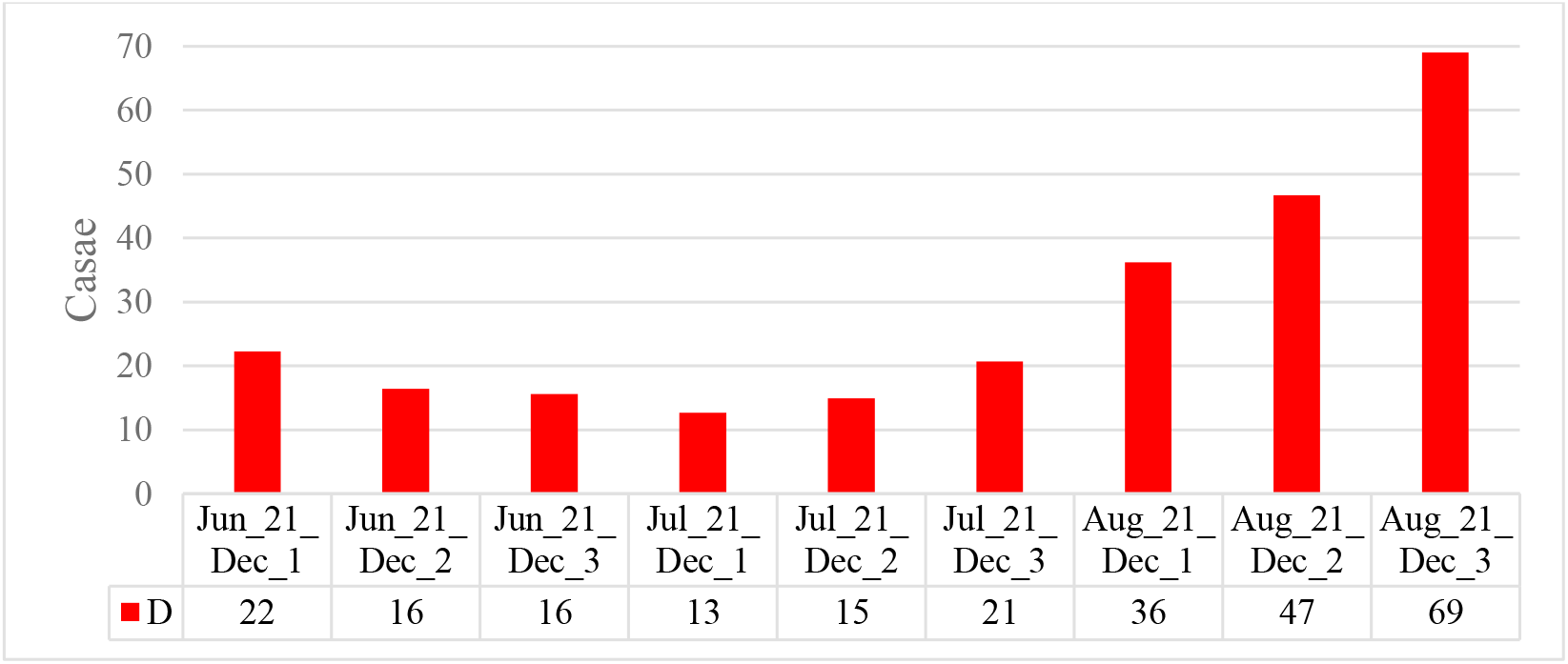
Mean values of deaths coronavirus-related cases in different decades of months in Georgia from June 2021 to August 2021.

Mean decade values of infection rate coronavirus-related cases (Fig. 10) varies from 2.51 % (2 Decade of June) to 10.88 % (2 Decade of August).

**Fig. 10.**
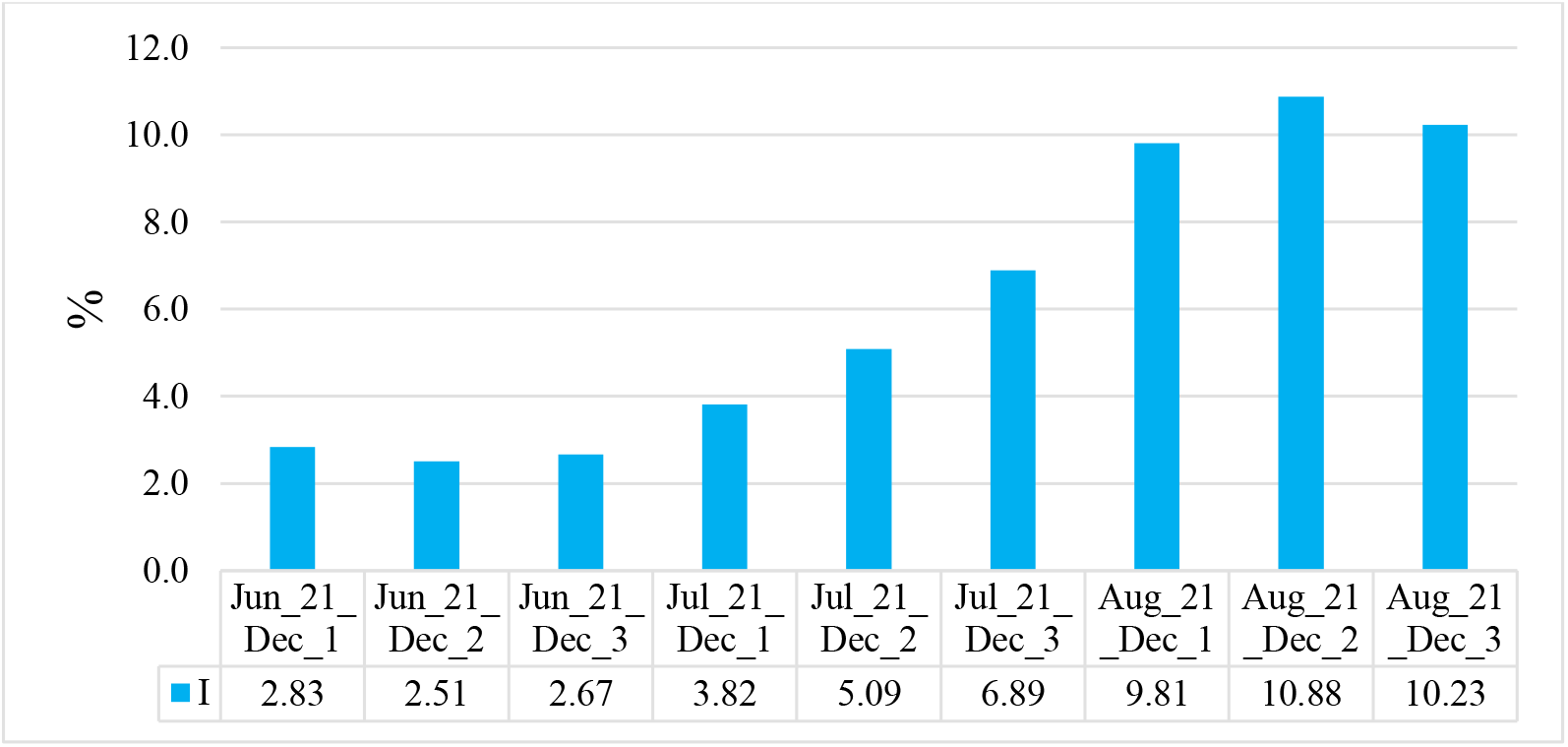
Mean values of infection rate coronavirus-related cases in different decades of months in Georgia from June 2021 to August 2021.

Time changeability of the daily values of C, R, D and I are satisfactorily described by the tenth order polynomial (Table 6, Fig. 11-13). For clarity, the data in Fig. 12 are presented in relative units (%) in relation to their average values.

**Table 6.**
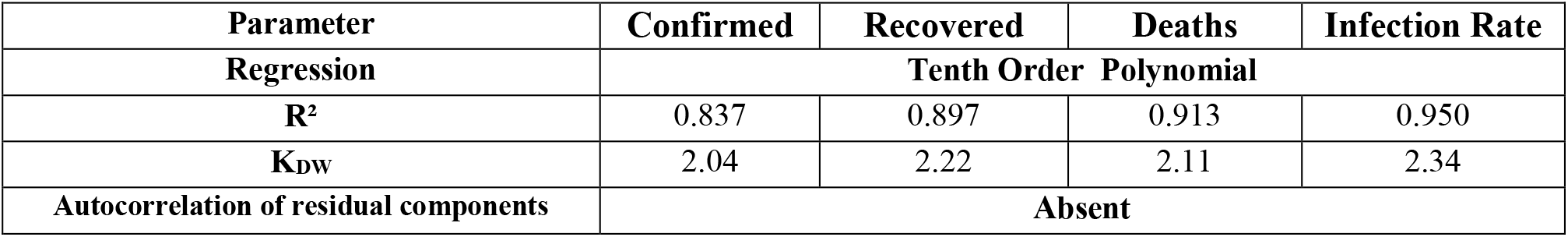
Form of the equations of the regression of the time changeability of the daily values of C, R and D from June 01, 2021 to August 31, 2021 in Georgia. The level of significance of R^2^ is not worse than 0.001.

| Parameter | Confirmed | Recovered | Deaths | Infection Rate |
| --- | --- | --- | --- | --- |
| Regression | Tenth Order Polynomial |  |  |  |
| $R^2$ | 0.837 | 0.897 | 0.913 | 0.950 |
| K<sub>DW</sub> | 2.04 | 2.22 | 2.11 | 2.34 |
| Autocorrelation of residual components | Absent |  |  |  |

**Fig. 11.**
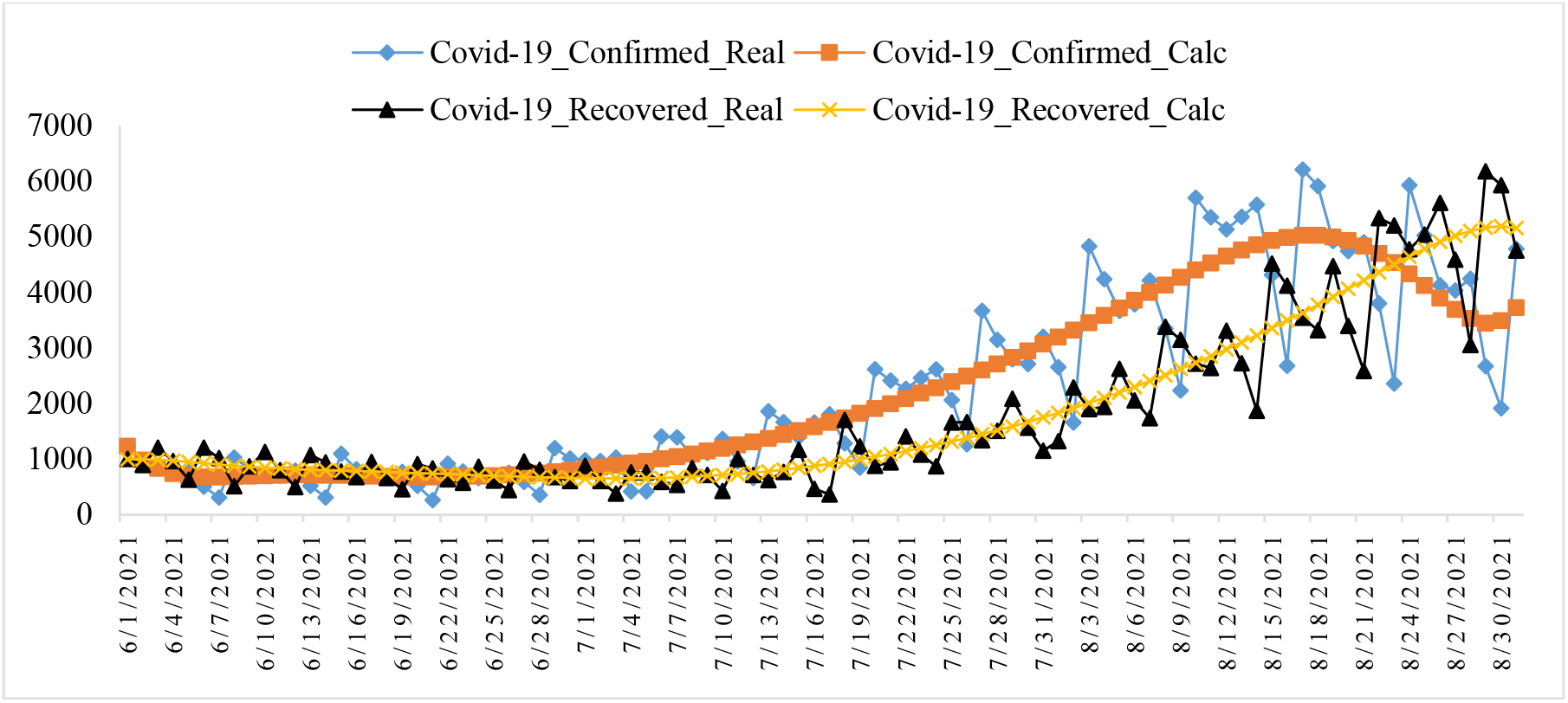
Changeability of the real and calculated daily values of coronavirus confirmed and recovered cases from June 01, 2021 to August 31, 2021 in Georgia.

**Fig. 12.**
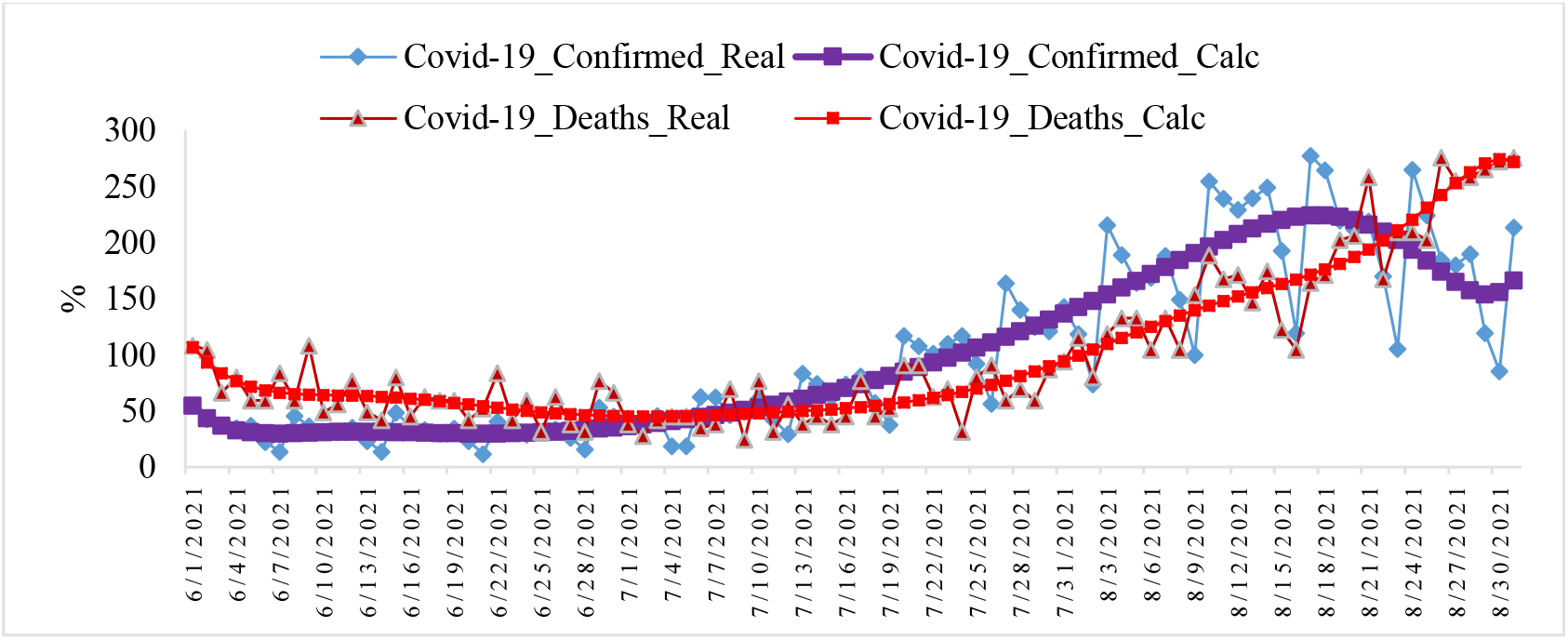
Changeability of the real and calculated daily values of coronavirus-related confirmed and deaths cases from June 01, 2021 to August 31, 2021 in Georgia (normed on mean values, %).

**Fig. 13.**
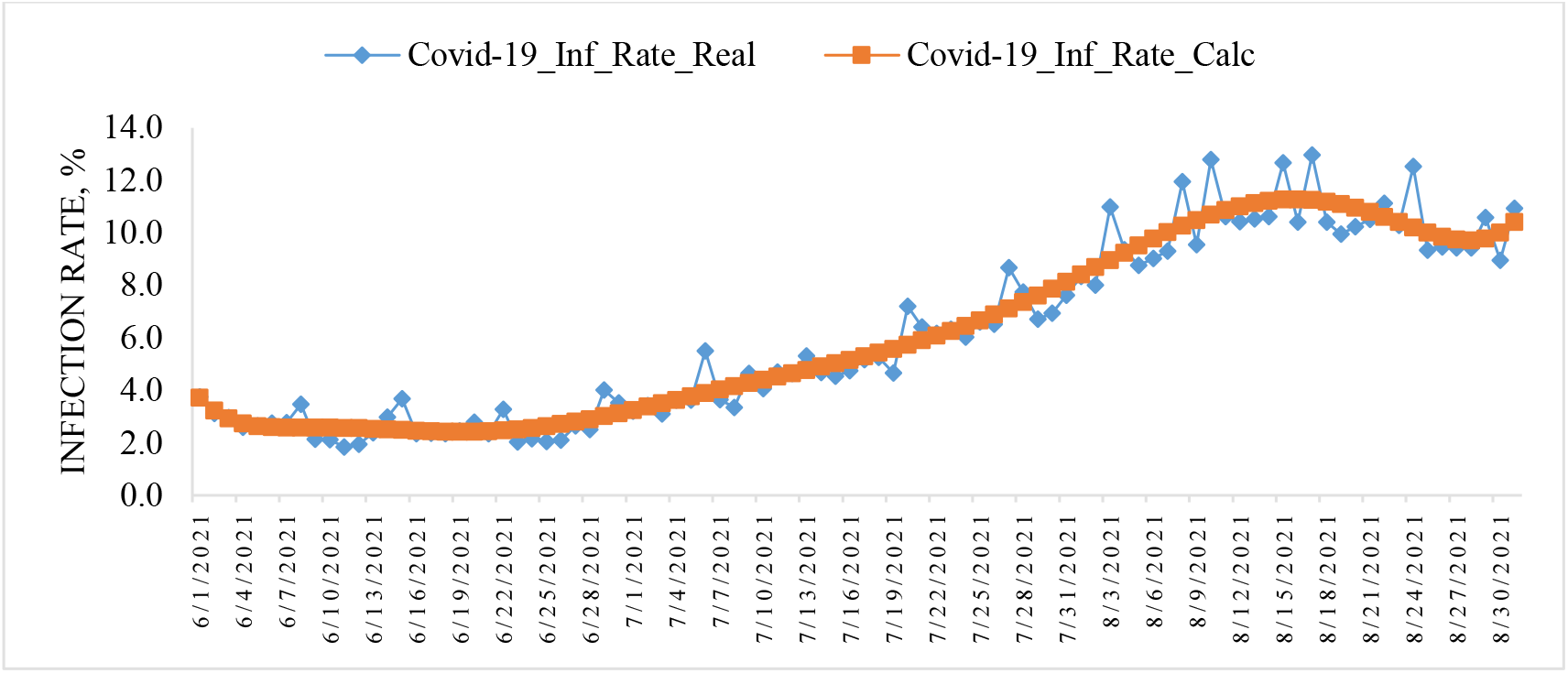
Changeability of the real and calculated daily values of coronavirus Infection Rate from June 01, 2021 to August 31, 2021 in Georgia.

Note that from Fig. 11 and 12, as in [7,8], clearly show the shift of the time series values of R and D in relation to C.

In Fig. 14-16 data about mean values of speed of change of confirmed, recovered, deaths coronavirus-related cases and infection rate in different decades of months in summer 2021 are presented.

**Fig. 14.**
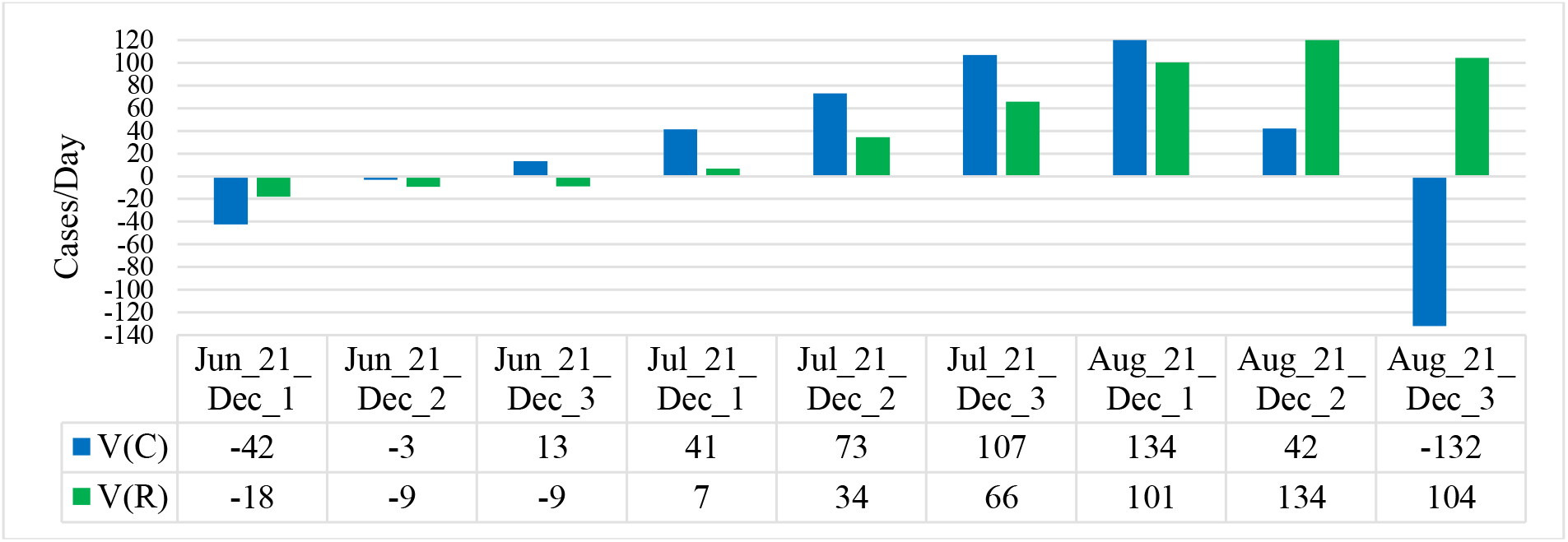
Mean values of speed of change of confirmed and recovered coronavirus-related cases in different decades of months in Georgia from June 2021 to August 2021.

**Fig. 15.**
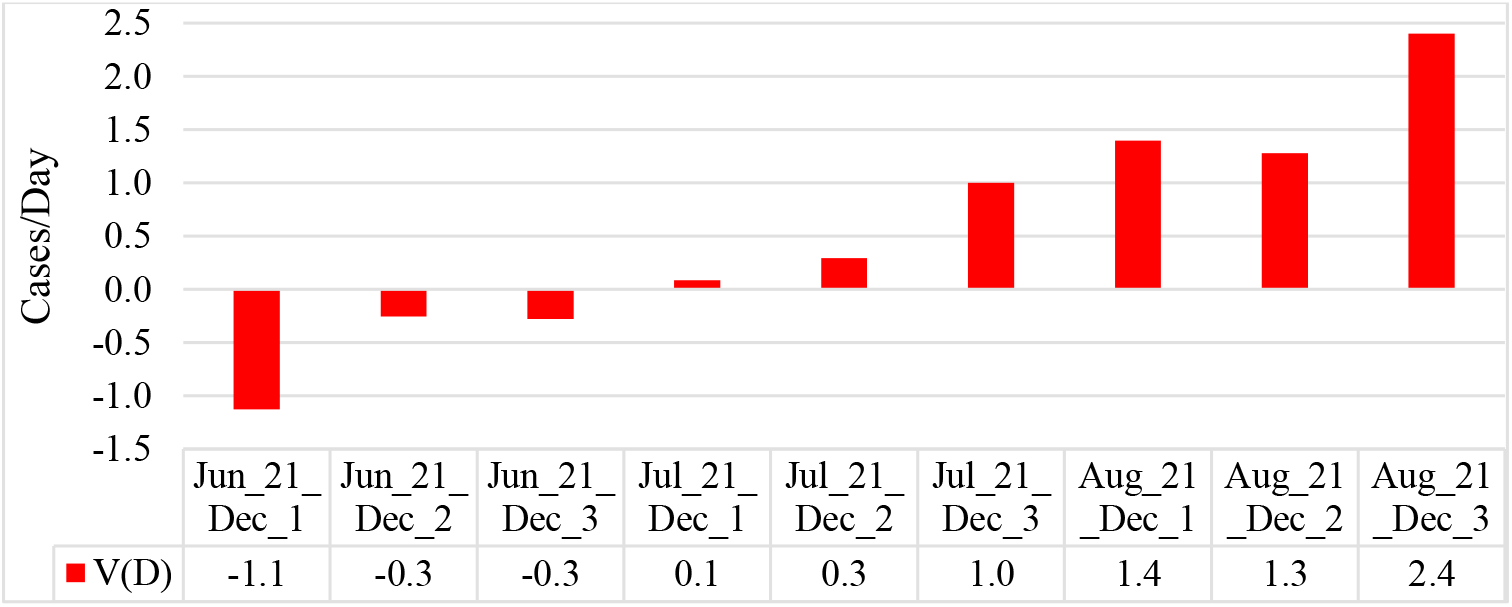
Mean values of speed of change of deaths coronavirus-related cases in different decades of months in Georgia from June 2021 to August 2021.

**Fig. 16.**
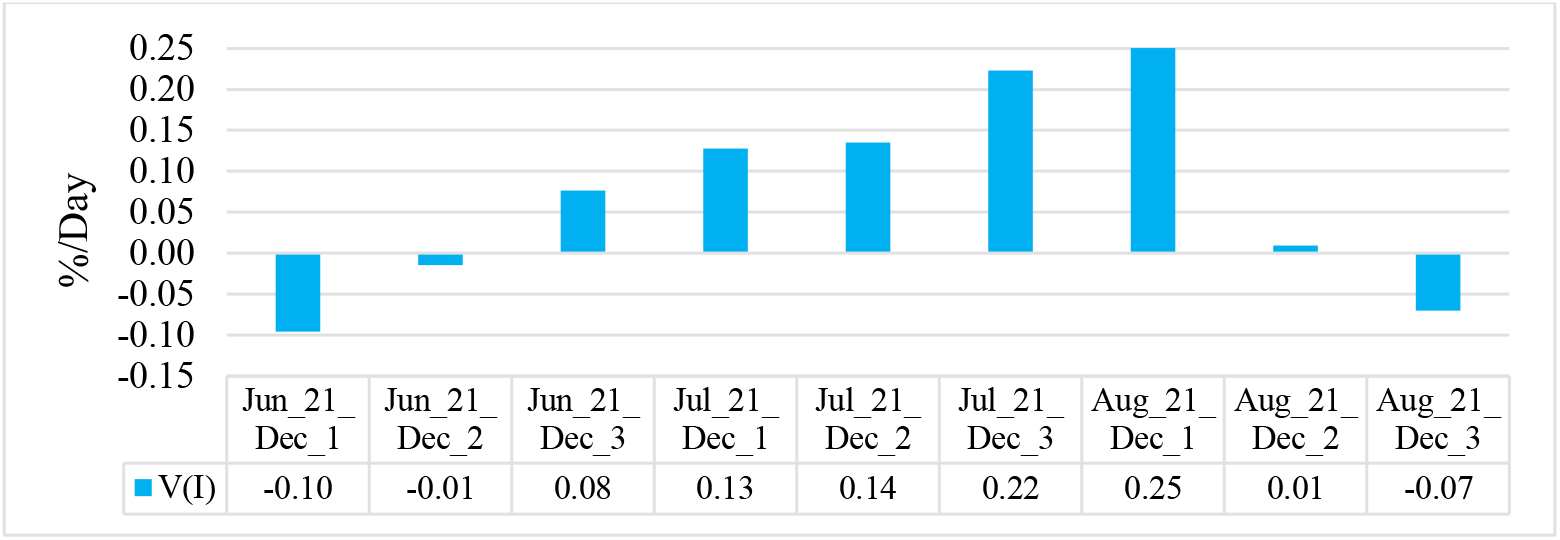
Mean values of speed of change of coronavirus infection rate in different decades of months in Georgia from June 2021 to August 2021.

Maximum mean decade values of investigation parameters are following: V(C) = +134 cases/day (1 Decade of August), V(R) = +134 cases/day (2 Decade of August), V(D) = +2.4 cases/day (3 Decade of August), V(I) = + 0.25 %/ day (1 decades of August). Min mean decade values of investigation parameters are following: V(C) = -132 cases/day (3 Decade of August), V(R) = -18 cases/day (1 Decade of June), V(D) = -1.1 cases/day (1 Decade of June), V(I) = -0.10 %/day (1 Decade of June).

Data about mean monthly values of C, R, D, I and its speed of change in Georgia in summer 2021 in Table 7 are presented. As follows from this Table there was an increase in average monthly values of C, R and I from June to August. Value of D decrease from June to July, and further increase to August.

**Table 7.** Mean monthly values of C, R, D, I and its speed of change in Georgia from June 2021 to August 2021.

| Parameter | C | R | D | I, % | V(C) | V(R) | V(D) | V(I) |
| --- | --- | --- | --- | --- | --- | --- | --- | --- |
| Jun_2021 | 737 | 804 | 18 | 2.67 | -9.6 | -11.9 | -0.5 | -0.01 |
| Jul_2021 | 1724 | 989 | 16 | 5.32 | 74.8 | 36.5 | 0.5 | 0.16 |
| Aug_2021 | 4202 | 3549 | 51 | 10.30 | 14.7 | 113.0 | 1.7 | 0.06 |

The values of V(C), V(R), V(D) and V(I) changes as follows: V(C) – from -9.6 cases/day (June) to +74.8 cases/day (July); V(R) - from -11.9 cases/day (June) to +113.0 cases/day (August); V(D) - from -0.5 cases/day (June) to +1.7 cases/day (August) and V(I) - from -0.01 %/day (June) to +0.16 %/day (July).

In Fig. 17 data about connection of 14-day moving average of deaths cases due to COVID-19 in Georgia with 14-day moving average of infection rate from December 18, 2020 until August 31, 2021 are presented. As follows from Fig. 17, in general, with an increase of the infection rate is observed increase of deaths cases. However, (Fig. 18), in August 2021 (I=10.4%), the average death rate from coronavirus (D=51.2) was more as in December 2020 (I=19.2, D= 40.0). This fact needs special analysis. It is possible that this is due to the predominance of the delta variant of the coronavirus in Georgia in the summer of 2021.

**Fig. 17.**
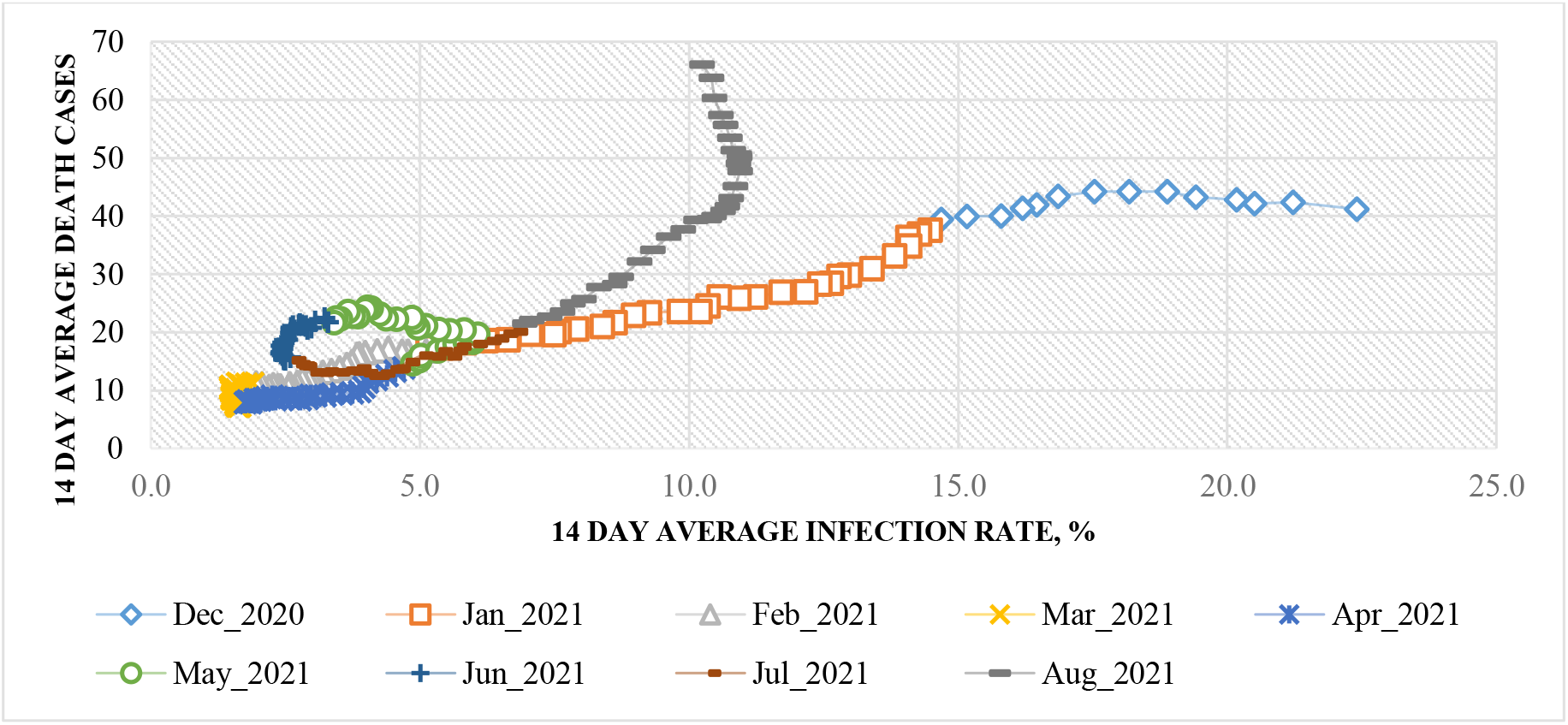
Connection of 14-day moving average of deaths cases due to COVID-19 in Georgia with 14-day moving average of infection rate from December 18, 2020 until August 31, 2021.

**Fig. 18.**
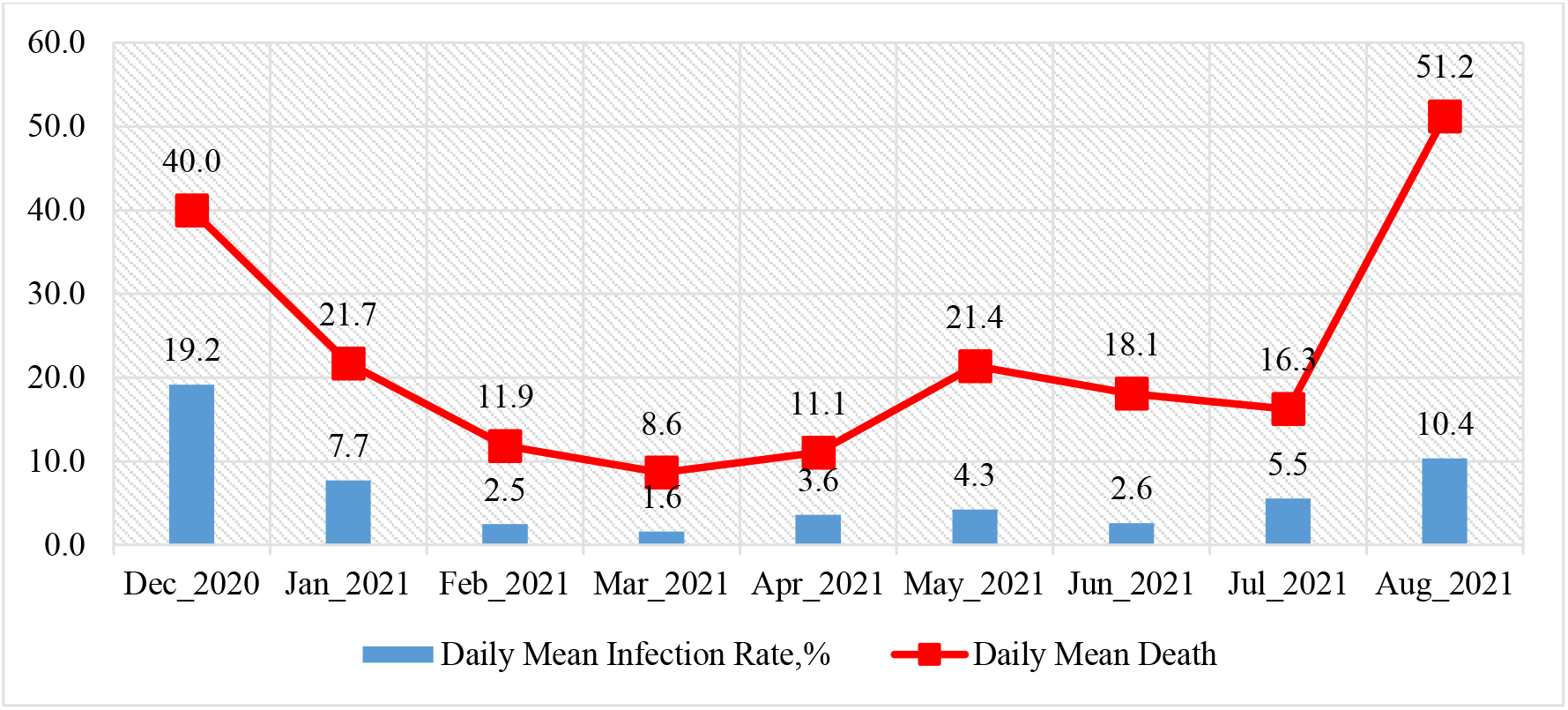
Mean monthly values of COVID-19 infection rate and deaths cases in Georgia from December 2020 to August 2021.

Using the data in Fig. 18, a linear regression graph between the monthly mean values of D and I is obtained (Fig. 19). As follows from Fig. 19, in general a high level of linear correlation between these parameters is observed. This Fig. also clearly demonstrates the anomaly high mortality from coronavirus in August 2021 with relatively low value of infection rate in comparison with December 2020, which reduces the level of correlation between D and I.

**Fig. 19.**
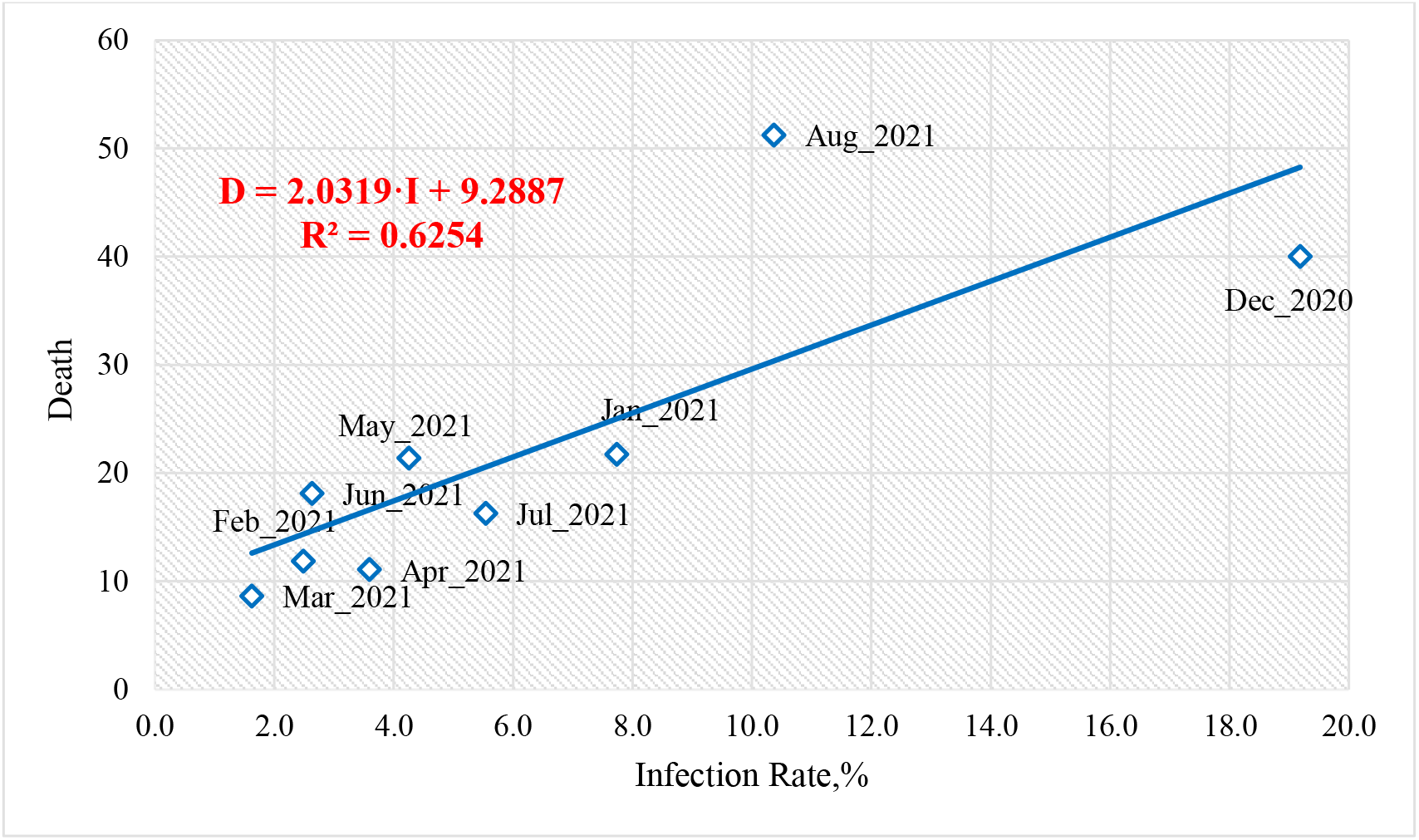
Linear correlation and regression between the mean monthly values of COVID-19 infection rate and deaths cases in Georgia from December 2020 to August 2021. R=0.79, α≈0.01

As noted in [7,8] and above (Fig. 11 and 12), there is some time-lag in the values of the time series R and D with respect to C. An estimate of the values of this time-lag is given below (Fig. 20).

**Fig. 20.**
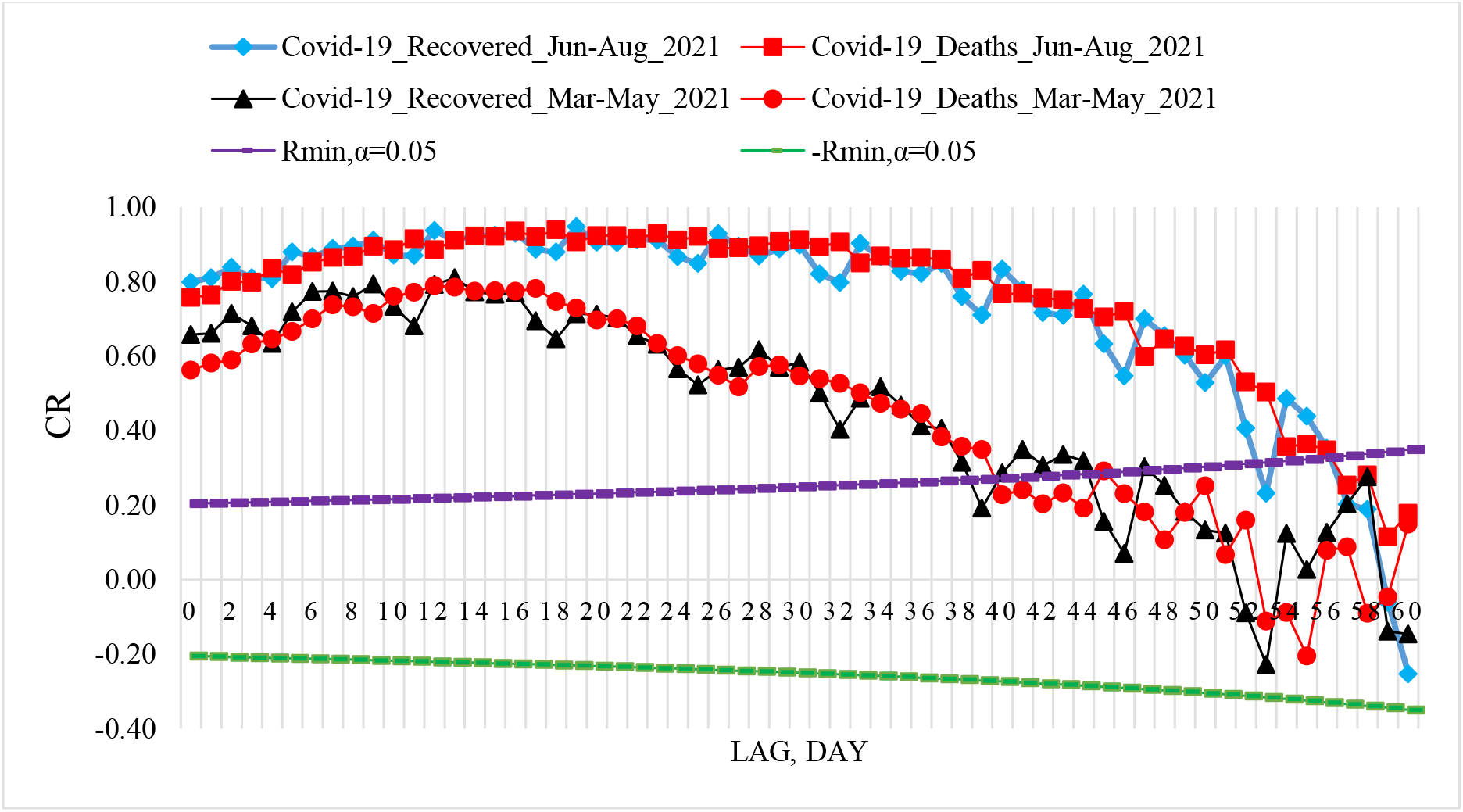
Coefficient of cross-correlations between confirmed COVID-19 cases (normed to tests number) with recovered and deaths cases in Georgia from 01.03.2021 to 31.05.2021 and from 01.06.2021 to 31.08.2021.

Cross-correlations analysis between confirmed COVID-19 cases with recovered and deaths cases shows, that in spring 2021 the maximum effect of recovery is observed 9 and 13 days after infection, and deaths - after 12-17 days (Fig. 20 and [8]). In spring 2021 the cross-correlation of the C values with the R and D values weakens completely 38-45 days after infection. Maximum values of RC were 0.81 for R, and 0.79 for D.

In summer 2021 the maximum effect of recovery is observed 19 days after infection (RC=0.95), and deaths - after 16 and 18 days (RC=0.94). In contrast to the spring of 2021, in the summer of 2021, a high level of cross-correlation of C values with R and D values to 40 days after infection is observed (RC≥ 0.80). In summer 2021 the cross-correlation of the C values with the R and D values weakens completely 56-58 days after infection. So, in Georgia in summer 2021, the duration of the impact of the delta variant of the coronavirus on people (recovery, mortality) could be up to two months.

### 3.4 Comparison of real and calculated prognostic daily and two weekly data related to the New Coronavirus COVID-19 pandemic in Georgia in summer 2021

In Fig. 21-24 and Table 8 examples of comparison of real and calculated prognostic daily and mean two weekly data related to the COVID-19 coronavirus pandemic in Georgia in summer 2021are presented. Note that the results of the analysis of two-week forecasting of the values of C, D and I, information about which was regularly sent to the National Center for Disease Control & Public Health of Georgia and posted on the Facebook page https://www.facebook.com/Avtandil1948/.

**Table 8.** Verification of decade and two-week interval prediction of COVID-19 confirmed infection cases, deaths cases and infection rate in Georgia from 01.06.2021 to 31.08.2021.

| Date | Low Level |  |  |  | Forecast<br>Central Point | Real Data | Forecast<br>Central Point | Upp Level |  |  |  |
| --- | --- | --- | --- | --- | --- | --- | --- | --- | --- | --- | --- |
|  | 99.99% | 99% | 95% | 67% |  |  |  | 67% | 95% | 99% | 99.99% |
|  | Noticeable, Significant, Sharp<br>Improvement of Pre-Forecast<br>State |  |  |  | Pre-Predicted State<br>Preservation |  | Pre-Predicted State<br>Preservation |  | Noticeable, Significant, Sharp<br>Deterioration of Pre-Forecast<br>State |  |  |
|  | COVID-19 Infection Cases |  |  |  |  |  |  |  |  |  |  |
| 6/01/2021-6/15/2021 | 5 | 107 | 245 | 511 | 798 | 760 | 798 | 1124 | 1454 | 1660 | 2101 |
| 6/16/2021-6/30/2020 | 3 | 70 | 174 | 448 | 715 | 714 | 715 | 1008 | 1304 | 1489 | 1884 |
| 7/01/2021-7/15/2021 | 0 | 24 | 145 | 392 | 691 | 1110 | 691 | 1000 | 1312 | 1507 | 1924 |
| 7/16/2021-7/31/2021 | 132 | 394 | 637 | 1047 | 1452 | 2300 | 1452 | 1857 | 2268 | 2524 | 3071 |
| 8/01/2021-8/15/2021 | 963 | 1598 | 2020 | 2726 | 3424 | 4136 | 3424 | 4121 | 4828 | 5269 | 6211 |
| 8/16/2021-8/31/2020 | 1724 | 3049 | 3673 | 4672 | 5658 | 4265 | 5658 | 6645 | 7644 | 8268 | 9600 |
|  | COVID-19 Death Cases |  |  |  |  |  |  |  |  |  |  |
| 6/01/2021-6/15/2021 | 0 | 4 | 7 | 12 | 17 | 21 | 17 | 23 | 28 | 31 | 38 |
| 6/16/2021-6/30/2020 | 0 | 2 | 6 | 13 | 20 | 16 | 20 | 27 | 34 | 38 | 47 |
| 7/01/2021-7/15/2021 | 0 | 0 | 0 | 6 | 15 | 12 | 15 | 24 | 33 | 38 | 50 |
| 7/16/2021-7/31/2021 | 0 | 0 | 0 | 3 | 12 | 20 | 12 | 20 | 29 | 34 | 45 |
| 8/01/2021-8/15/2021 | 0 | 9 | 13 | 20 | 27 | 39 | 27 | 34 | 41 | 45 | 54 |
| 8/16/2021-8/31/2020 | 5 | 22 | 26 | 37 | 49 | 63 | 49 | 60 | 71 | 77 | 93 |
|  | COVID-19 Infection Rate, % |  |  |  |  |  |  |  |  |  |  |
| 6/01/2021-6/15/2021 | 0.0 | 0.1 | 0.4 | 1.2 | 2.8 | 2.7 | 2.8 | 4.4 | 6.1 | 7.1 | 9.3 |
| 6/16/2021-6/30/2020 | 0.1 | 0.3 | 0.5 | 0.9 | 2.4 | 2.6 | 2.4 | 3.8 | 5.3 | 6.2 | 8.1 |
| 7/01/2021-7/15/2021 | 0.2 | 0.8 | 1.4 | 2.4 | 3.3 | 4.1 | 3.3 | 4.3 | 5.3 | 5.9 | 7.2 |
| 7/16/2021-7/31/2021 | 0.9 | 2.0 | 2.9 | 4.2 | 5.6 | 6.4 | 5.6 | 6.9 | 8.3 | 9.2 | 11.0 |
| 8/01/2021-8/15/2021 | 5.3 | 6.5 | 7.1 | 8.0 | 8.9 | 10.2 | 8.9 | 9.9 | 10.8 | 11.4 | 12.6 |
| 8/16/2021-8/31/2020 | 7.2 | 9.6 | 10.8 | 12.6 | 14.4 | 10.4 | 14.4 | 16.2 | 18.0 | 19.1 | 21.6 |

**Fig. 21.**
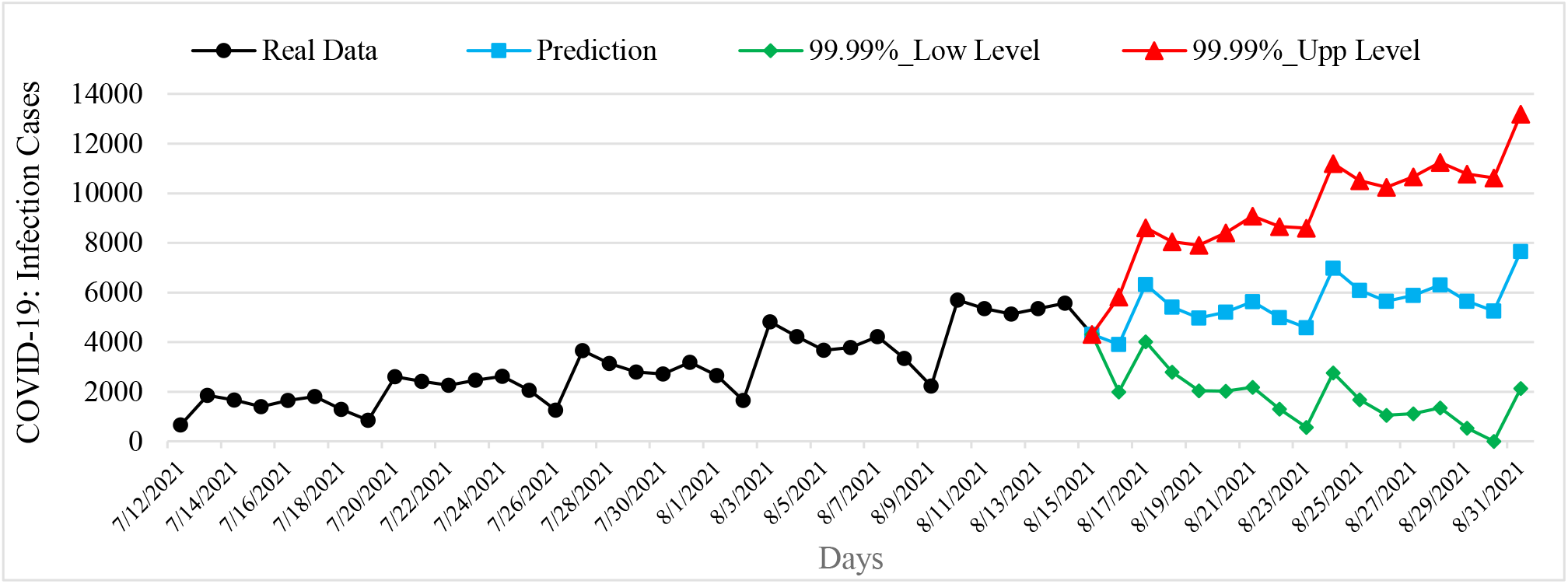
Example of Interval Prediction of COVID-19 Infection Cases in Georgia from 16.08.2021 to 31.08.2021.

**Fig. 22.**
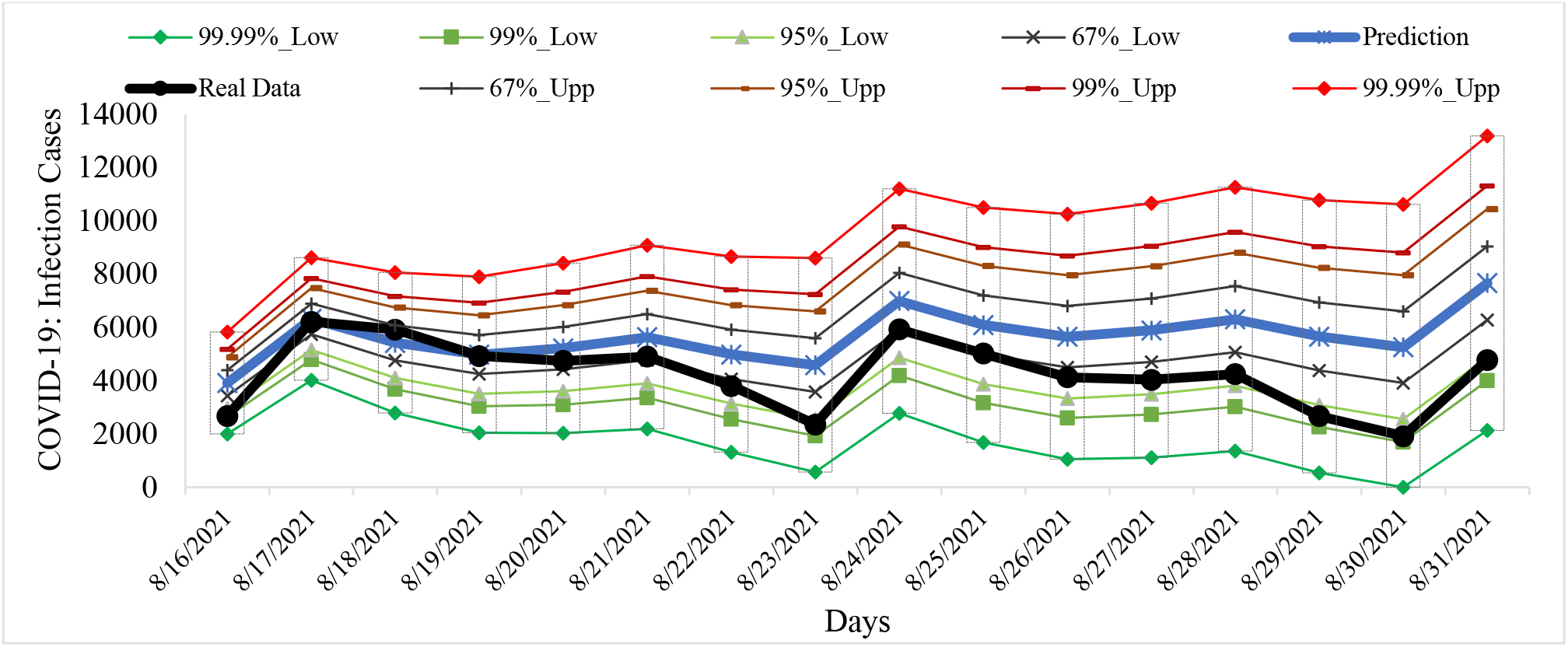
Example of Verification of Interval Prediction of COVID-19 Infection Cases in Georgia from 16.08.2021 to 31.08.2021.

**Fig. 23.**
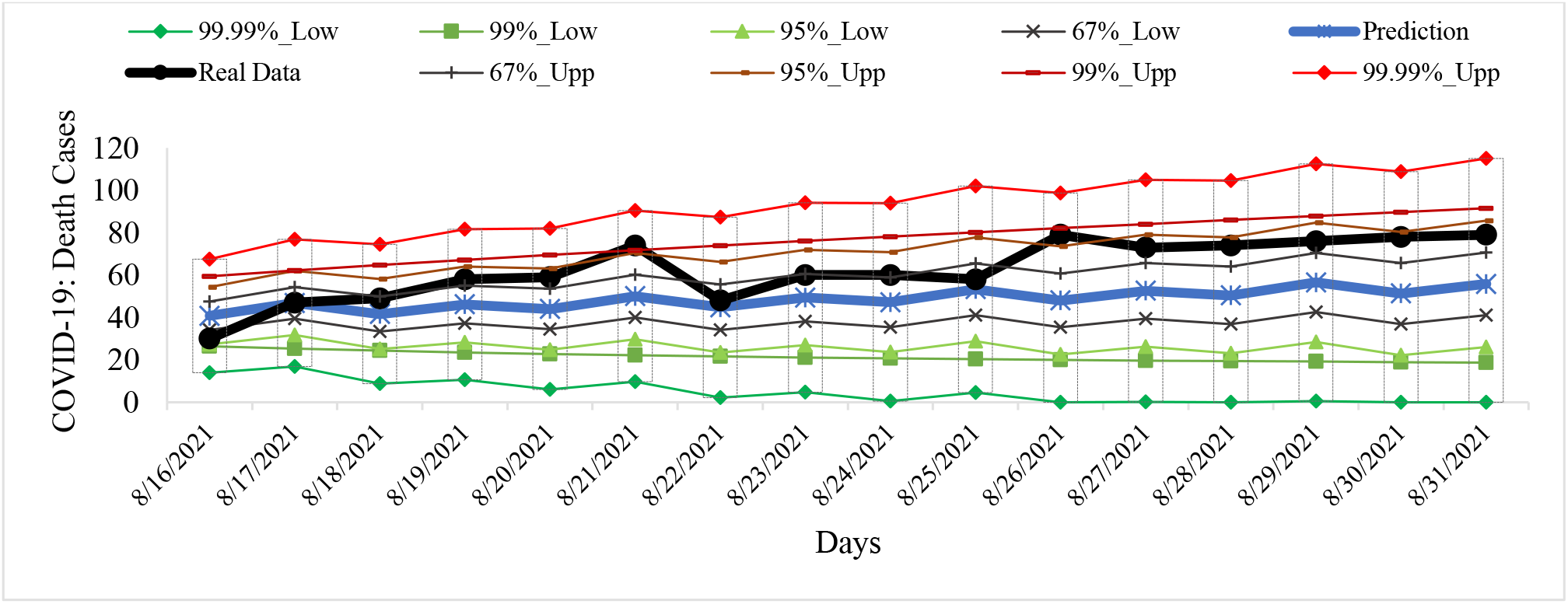
Example of Verification Interval Prediction of COVID-19 Death Cases in Georgia from 16.08.2021 to 31.08.2021.

**Fig. 24.**
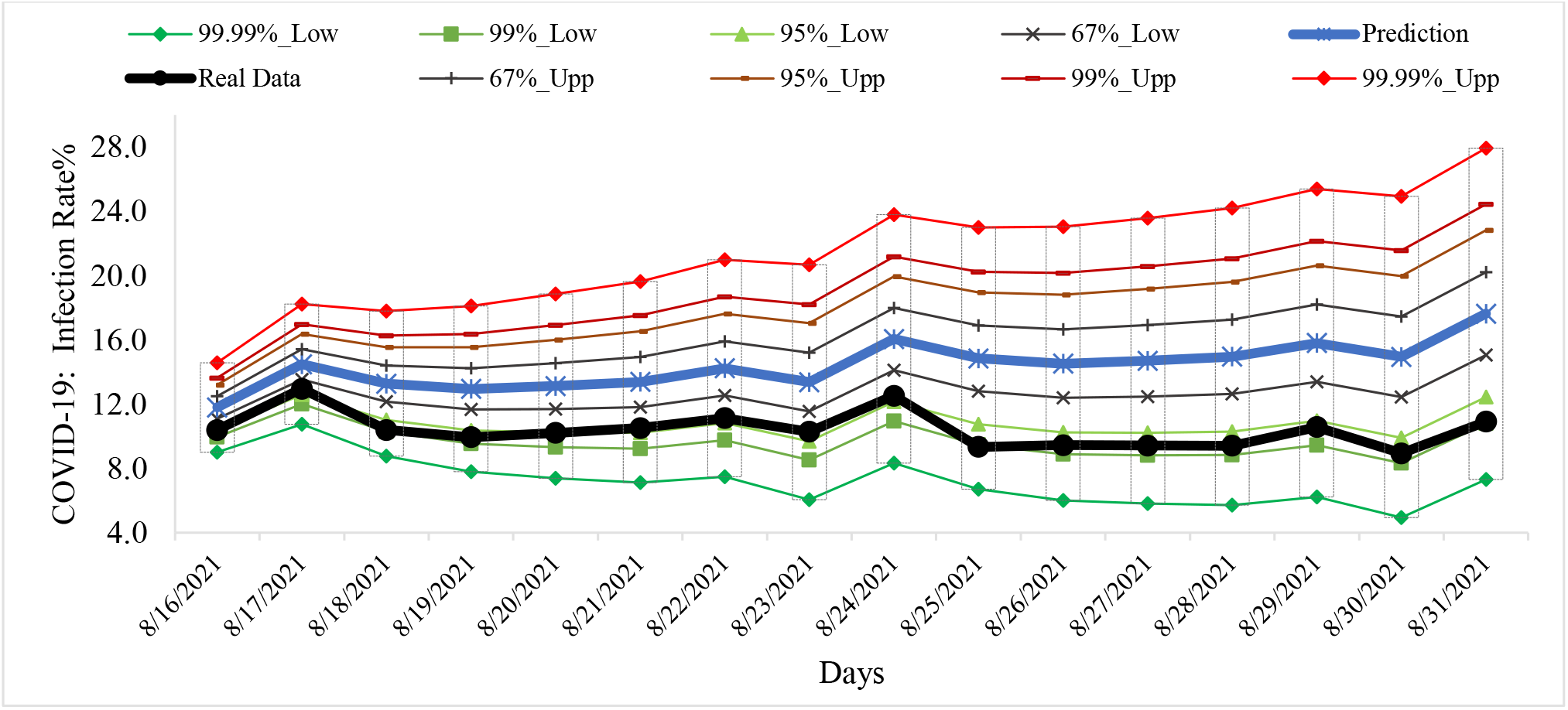
Example of Verification Interval Prediction of COVID-19 Infection Rate in Georgia from 16.08.2021 to 31.08.2021.

Comparison of real and calculated predictions data of C, D and I shown, that two-week daily and mean two-week real values of C, D and I practically falls into the 67% - 99.99% confidence interval of these predicted values for the specified time periods (Fig. 21-24, data from https://www.facebook.com/Avtandil1948/, Table 8).

For all forecast periods (6 forecasting cases), the stability of real time series of observations of these parameters (period for calculating the forecast + forecast period) remained (Table 9).

**Table 9.** Change in the forecast state of C, D and I in relation to the pre-predicted one according to Table 1 scale [6].

| Date | COVID-19 Infection Cases |
| --- | --- |
| 6/01/2021- 6/15/2021 | Preservation, stability of a time-series of observations |
| 6/16/2021-6/30/2020 | Preservation, stability of a time-series of observations |
| 7/01/2021-7/15/2021 | Noticeable deterioration, stability of a time-series of observations |
| 7/16/2021-7/31/2021 | Significant deterioration, stability of a time-series of observations |
| 8/01/2021-8/15/2021 | Noticeable deterioration, stability of a time-series of observations |
| 8/16/2021-8/31/2020 | Noticeable improvement, stability of a time-series of observations |
|  | **COVID-19 Death Cases** |
| 6/01/2021- 6/15/2021 | Preservation, stability of a time-series of observations |
| 6/16/2021-6/30/2020 | Preservation, stability of a time-series of observations |
| 7/01/2021-7/15/2021 | Preservation, stability of a time-series of observations |
| 7/16/2021-7/31/2021 | Preservation, stability of a time-series of observations |
| 8/01/2021-8/15/2021 | Noticeable deterioration, stability of a time-series of observations |
| 8/16/2021-8/31/2020 | Noticeable deterioration, stability of a time-series of observations |
|  | **COVID-19 Infection Rate** |
| 6/01/2021- 6/15/2021 | Preservation, stability of a time-series of observations |
| 6/16/2021-6/30/2020 | Preservation, stability of a time-series of observations |
| 7/01/2021-7/15/2021 | Preservation, stability of a time-series of observations |
| 7/16/2021-7/31/2021 | Preservation, stability of a time-series of observations |
| 8/01/2021-8/15/2021 | Noticeable deterioration, stability of a time-series of observations |
| 8/16/2021-8/31/2020 | Significant improvement, stability of a time-series of observations |

Thus, the daily two-week and mean two-week forecasted values of C, D and I quite adequately describe the temporal changes in their real values.

With September 1, 2021, we started monthly forecasting of C, D and I values.

An interval forecast check of confirmed COVID-19 cases, deaths and infection rates in Georgia from 01.09.2021 to 08.09.2021 confirms the representativeness of the monthly forecast for the specified time period [https://www.facebook.com/Avtandil1948/]. Further monitoring will determine the feasibility of this monthly forecast.

## Conclusion

In the future, it is planned to continue regular similar studies for Georgia in comparison with neighboring and other countries, in particular, checking the temporal representativeness of monthly interval statistical forecasting of the new coronavirus infection COVID-19, taking into account data on the periodicity, clarification of connections between various parameters of this infection, the influence of bioclimatic factors on this infection, etc.

## Data Availability

https://www.facebook.com/Avtandil1948/

## Notes

Source(s) of support: Nil.

Conflicting Interest – No.

### Competing Interest Statement

The authors have declared no competing interest.

### Clinical Trial

This article is a preprint and has not been certified by peer review [what does this mean?]. It reports new medical research that has yet to be evaluated and so should not be used to guide clinical practice.

### Clinical Protocols

https://www.facebook.com/Avtandil1948/

### Funding Statement

Budget financing

### Author Declarations

https://www.soothsawyer.com/john-hopkins-time-series-data-with-us-state-and-county-city-detail-historical/; https://data.humdata.org/dataset/total-covid-19-tests-performed-by-country https://stopcov.ge

